# Multi-omic profiling of cutaneous leishmaniasis infections reveals microbiota-driven mechanisms underlying disease severity

**DOI:** 10.1101/2023.02.02.23285247

**Authors:** Camila Farias Amorim, Victoria M. Lovins, Tej Pratap Singh, Fernanda O. Novais, Jordan C. Harris, Alexsandro S. Lago, Lucas P. Carvalho, Edgar M. Carvalho, Daniel P. Beiting, Phillip Scott, Elizabeth A. Grice

## Abstract

*Leishmania braziliensis* infection results in inflammation and skin injury, with highly variable and unpredictable clinical outcomes. Here, we investigated the potential impact of microbiota on infection-induced inflammatory responses and disease resolution by conducting an integrated analysis of the skin microbiome and host transcriptome on a cohort of 62 *L. braziliensis*-infected patients. We found that overall bacterial burden and microbiome configurations dominated with *Staphylococcus* spp. were associated with delayed healing and enhanced inflammatory responses, especially by IL-1 family members. Dual RNA-seq of human lesions revealed that high lesional *S. aureus* transcript abundance was associated with delayed healing and increased expression of IL-1β. This cytokine was critical for modulating disease outcome in *L. braziliensis*-infected mice colonized with *S. aureus*, as its neutralization reduced pathology and inflammation. These results implicate the microbiome in cutaneous leishmaniasis disease outcomes in humans and suggest host-directed therapies to mitigate the inflammatory consequences.

## INTRODUCTION

Cutaneous leishmaniasis is a neglected tropical disease caused by protozoan parasites transmitted by sand flies. The clinical manifestation of leishmania infection is highly variable, ranging from single healing skin lesions to severe chronic ulcers, including disseminated and mucosal manifestations, all of which can be disfiguring ^1–4^. The parasite burden and the host immune response can influence where a patient falls on this clinical spectrum. Notably, some of the most severe forms of the disease are characterized by chronic inflammation despite of parasite control ^2^. Since parasite-targeted drug treatment such as pentavalent antimony (Sb^v^) is associated with high rate of failure in our endemic area and there is no vaccine for the disease, identifying mechanisms driving destructive immunopathologic responses could provide additional therapeutic targets to ameliorate the more severe forms of cutaneous leishmaniasis.

The skin microbiome contributes to homeostatic mechanisms that fortify the skin’s barrier function ^5^. In contrast, while a breach of the skin barrier, such as that caused by leishmania infection, disturbs the commensal microbiota and exposes underlying tissues to invasion by microbes ^6^. Several studies suggest that the skin microbiome promotes increased disease in experimental models of cutaneous leishmaniasis ^7–9^. For example, germ-free (GF) mice infected with *Leishmania major* develop smaller lesions than those observed in conventional mice, and colonization of germ-free mice with *S. epidermidis* restores ulcer development in *L. major* infected mice ^8^. In conventionally raised mice, colonization or co-infection with *S. aureus, S. epidermidis*, or *S. xylosus* promotes increased lesional pathology, accompanied by increased inflammation and higher expression of pro-inflammatory cytokines such as IL-1β and IL-17 in lesions compared to infection with *L. major* alone ^7,9,10^. While these experimental models suggest a pathological role for the microbiota, the relevance of these findings to the clinical course of human *Leishmania* infection and lesion resolution remains unclear.

To address this gap, we conducted an integrative multi-omics meta-analysis in a cohort of *L. braziliensis* patients. We performed 16S rRNA amplicon sequencing (16S-seq) of lesional and contralateral skin swabs to profile the microbial communities longitudinally. In parallel, lesional biopsies were collected for RNA-sequencing (RNA-seq) to profile the host transcriptome and quantify total bacterial burden. Bacterial burden was increased in lesions compared to intact, contralateral skin, and lesional burden was correlated with pro-inflammatory host gene expression and delayed healing. Lesions were grouped into 8 distinct clusters based on 16S-seq profiles, but the majority fell within a cluster defined by the high relative abundance of the genus *Staphylococcus*. We next constructed a custom *S. aureus* pangenome using clinical isolates and quantified *S. aureus* reads detected in each lesional biopsy. High *S. aureus* read counts were associated with increased expression of cytolytic-encoding genes and IL-1-related gene transcription. Importantly, we found delayed healing in patients with high *S. aureus* read counts. Finally, to determine if *S. aureus*-induced IL-1β contributes to increased disease, we neutralized IL-1β signaling in *L. braziliensis* infected mice colonized with a *S. aureus* clinical isolate. Our findings confirmed that colonization with *S. aureus* promotes increased lesion size and pathology in an IL-1β-dependent manner. These findings demonstrate the significant impact of the skin microbiome on human leishmanial infections and highlight the potential of implementing microbiota-directed therapeutic strategies in this disease to improve clinical outcomes.

## RESULTS

### L. braziliensis-infected patient cohort and associated datasets

To investigate whether the microbiota influences skin inflammatory immune responses and clinical outcomes in patients infected with *L. braziliensis*, we carried out an integrative analysis of a multi-omics dataset. Samples collected from a cohort of 62 subjects infected with *L. braziliensis* were used to generate the following datasets: This dataset was composed of 1) a total transcriptome RNA-seq dataset from 51 lesion biopsies collected prior to pentavalent antimony (Sb^v^) treatment; 2) a 16S-seq dataset of swabs collected from the surface of the lesion prior to Sb^v^ treatment and on follow up visits for a subset of patients (visit days 30-240); 3) a library of *Staphylococcus aureus* isolates cultured from the lesions and their genome sequences; and 4) clinical metadata with patient demographics. The cohort consisted of 20 females and 42 males, with an average age of 30 years (range 17-56 years). Clinical metadata collected included lesion size and location, delayed type hypersensitive (DTH) measurements, lymphadenopathy, time to heal, and treatment outcome. In most cases, multiple sample types were collected from the same patient. The study design and inventory are detailed in Supplemental Fig. 1, Supplemental Table 1 and Supplemental Table 2).

### The lesional bacterial burden is associated with enhanced host inflammatory gene expression

To investigate the role of the skin microbiome in mediating host transcriptional pathways in *L. braziliensis* infections, we extracted RNA and made cDNA libraries from biopsies of leishmanial lesions and control skin of uninfected individuals. In parallel with RNA-seq analysis, we used the same cDNA libraries to estimate lesional bacterial burden by quantitative PCR targeting of the prokaryotic 16S ribosomal RNA (rRNA) gene (Fig. 1A) and found that the bacterial burden was elevated in lesions compared to contralateral healthy skin, P<0.01 (Fig. 1B). We performed a continuous Differential Gene Expression (DGE) analysis and identified 148 host genes that positively correlated with the lesional bacterial burden (linear model slope coefficient>0.2 and P<0.01) (Supplemental Table 3). Gene Ontology (GO) analysis of this gene list revealed enrichment for pro-inflammatory immune responses, with a significant participation of genes encoding for members of the IL-1 signaling pathway (*IL1A, IL1B, IL1RN* and *IL1R2*) and neutrophil chemotaxis, represented mainly by the CXCL-family of chemotactic encoding genes (Fig. 1C and 1D, Supplemental Table 4).

**Fig. 1.**
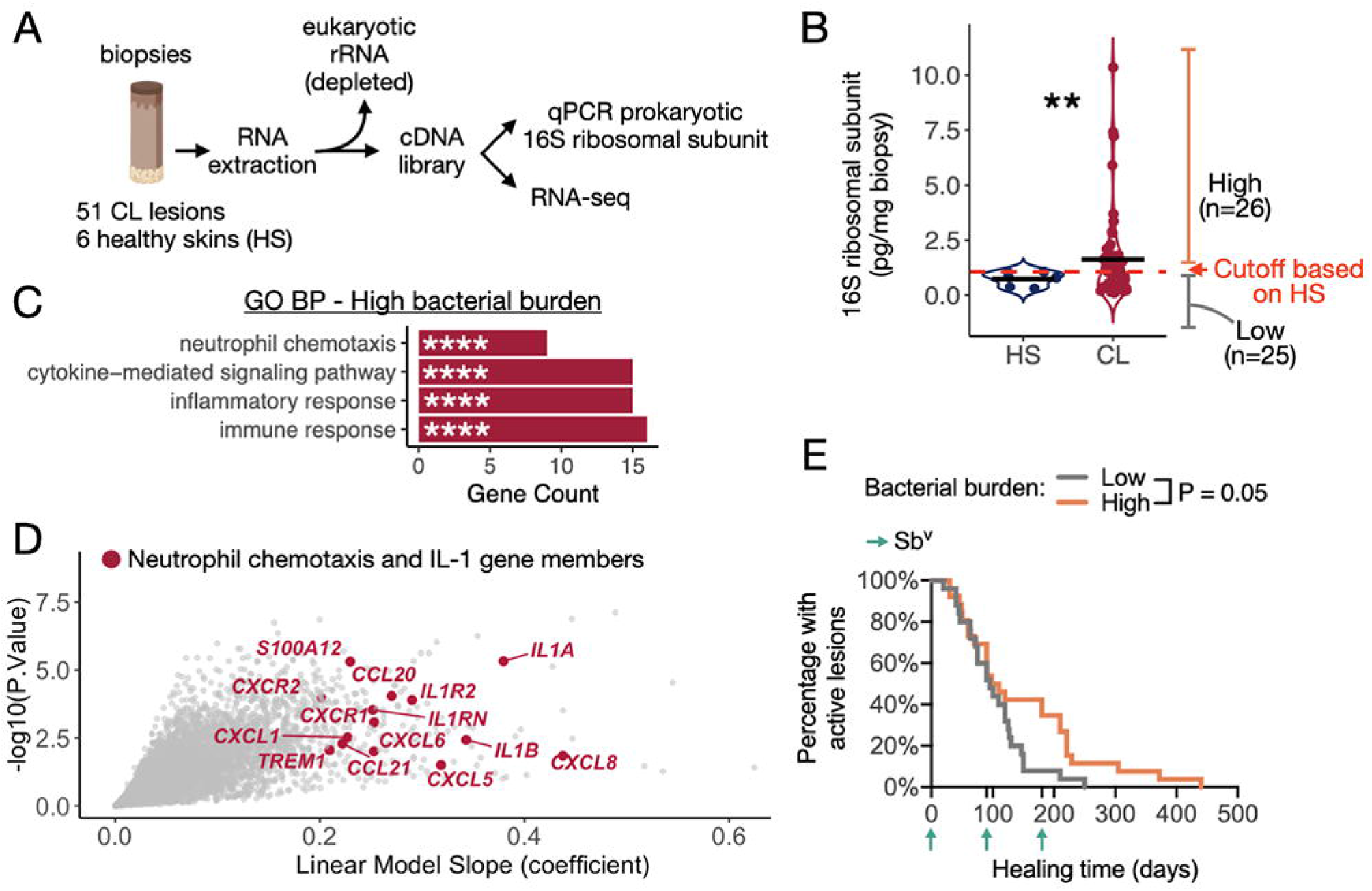
Lesional bacterial burden is associated with enhanced pro-inflammatory gene expression and delayed healing time. A) Schematics explaining the use of cDNA libraries from punch biopsies to perform RNA-seq and qPCR targeting the prokaryotic 16S rRNA gene to quantify total bacterial burden. Extracted RNA was depleted of eukaryotic rRNA prior to cDNA conversion. B) Bacterial burden quantified by qPCR in biopsies collected from CL lesions and healthy skin (HS) control samples not infected by *L. braziliensis*. The mean for each group is represented, and a T-test was used to calculate the statistical significance **P<0.01. C and D) Continuous Differential Gene Expression (DGE) analysis performed on the bacterial burden variable, calculated by the R *limma* package. GO analysis accessing Biological Processes (BP) was performed on the list of DEGs positively correlated with the bacterial burden at the threshold of coefficient slope >0.2 and P<0.01, and the top statistically enriched genesets are represented with gene count in a bar plot. The scatter plot highlights the genes annotated as *Neutrophil chemotaxis* and genes encoding for members of the IL-1 signaling. E) Survival curve for lesion healing time comparing lesions with high and low bacterial burden. The threshold for dividing high and low bacterial burden lesions was based on the qPCR levels detected in HS biopsies. Log-rank (Mantel-Cox) test was used to calculate the statistical significance. GO, Gene Ontology; ES, enrichment score. DEGs, differentially expressed genes.

As our results suggest that high levels of bacteria in the lesions drive the expression of pro-inflammatory gene expression programs, we hypothesized that bacterial burden would contribute to poor clinical outcomes in *L. braziliensis* patients. Based on bacterial 16S rRNA transcript levels in healthy contralateral skin, we split the cutaneous leishmaniasis cohort into high and low bacterial burden groups (n=26 and n=25 subjects, respectively) and compared healing times. Subjects with a high bacterial burden exhibited a delayed response to therapy (P = 0.05), with some lesions persisting >300 days (Fig. 1E). These findings motivated further investigation into the lesional microbiota and its potential impact on the immunopathology associated with cutaneous leishmaniasis.

### Staphylococcus often dominates the community structure of the lesional microbiota

To identify the bacteria that might be involved with delayed lesion resolution, we profiled microbiota of swab specimens collected from lesions and contralateral skin of *L. braziliensis* patients before to the start of therapy using 16S rRNA amplicon sequencing (Fig. 2A). We found that the skin microbiome was dramatically affected by infection with *L. braziliensis* relative to contralateral skin, with bacterial alpha diversity significantly lower in lesions as measured by the Shannon diversity index (Fig. 2B). Microbial community structure was also affected by *L. braziliensis* infection relative to contralateral skin, as calculated by the Weighted UniFrac dissimilarity metrics (Fig. 2C). To identify bacterial taxa enriched in lesions at baseline (before to treatment), we performed differential taxa abundance analysis between the lesion and contralateral skin swabs. The genus *Staphylococcus* was the most differentially abundant genus in lesions, followed by *Corynebacterium* and *Streptococcus* (FC>2 and FDR<0.05) (Fig. 2D). We also captured a signature of microbial communities present in the contralateral uninfected skin of the patients and identified the top five differentially abundant genera as *Nesterenkonia, Cutibacterium, Kocuria, Micrococcus*, and *Brachybacterium* (Fig. 2D). These genera are from the families *Propionibacteriaceae, Micrococciaceae*, and *Dermabacteraceae* and are commonly found on human skin in varying abundances ^11^.

**Fig. 2.**
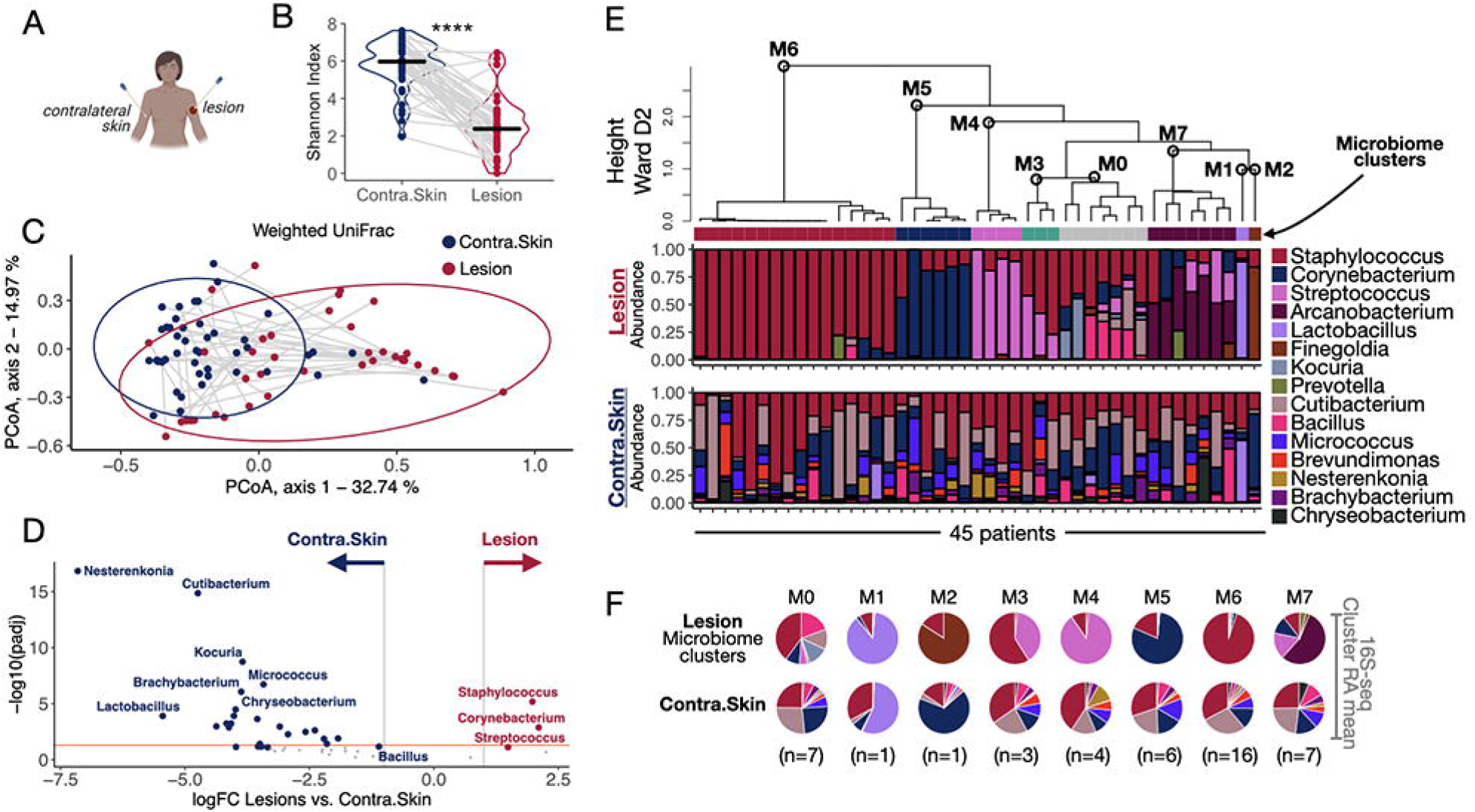
L. braziliensis infected lesions present distinct microbiome profiles. A) Swabs were collected from the lesion and contralateral skin sites for 16S-seq analysis. B) Shannon Index assessed bacterial alpha diversity. Gray lines connect the lesion and contralateral skin samples collected from the same patient. Paired t-test was used to calculate the statistical significance, ****P<0.0001. C) PCoA calculated with a weighted UniFrac dissimilarity analysis showing lesion and contralateral skin microbiome profiles placed in coordinates 1 and 2. D) Differential taxa abundance analysis between all the lesion and contralateral skin samples collected on Day 0. The orange line indicates adj. P. value = 0.05. E) Unsupervised HC calculated with Ward D2 agglomeration method clustered the lesion samples collected at Day 0 by their top 10 genera microbiome profiles (clusters M0-M7). The relative abundances of the top 10 taxa in the lesions are represented as stacked bar plots (top). The relative abundances associated with the microbiomes in the contralateral skin are represented right underneath the lesion sample from the same patient (bottom). F) The RAmeans of each taxon in lesions and contralateral skin samples per microbiome cluster (M0-M7) was calculated and visualized as a pie chart. PCoA, Principal coordinate analysis; HC, Hierarchical clustering; RAmean, Relative abundance mean.

To further investigate lesional bacterial communities, we reduced the dimensionality of the 16S-seq dataset to obtain the ten most abundant taxa. Unsupervised hierarchical clustering (HC) analysis based on the relative abundance of these taxa revealed eight distinct microbiome profiles, or “clusters” (Fig. 2E and 2F). The thresholds that defined these clusters were based on a consensus of relative abundance mean per sample (Supplemental Fig. 2A), and number of dominant taxa in the community (>80% of total dominance and >2 taxa, respectively). The largest cluster, microbiome cluster 6 (M6), included 16 patients with lesions presenting dominant colonization by *Staphylococcus* (95% relative abundance mean, RAmean). This was followed by clusters: M7, with seven patients presenting unique colonization by *Arcanobacterium* (56% RAmean), *Streptococcus* (17% RAmean), *Corynebacterium* (11% RAmean), and *Staphylococcus* (11% RAmean); M5 with six patients presenting a *Corynebacterium* dominant colonization (80% RAmean); M4 with four patients with a *Streptococcus* dominant colonization (90% RAmean), M3 with three patients presenting a co-colonization by *Streptococcus* and *Staphylococcus* (40% and 60% RAmeans, respectively), and two patients with either *Lactobacillus* or *Finegoldia* dominant colonization (M1 88% and M2 85% RAmean, respectively) (Fig. 2E and 2F). Seven patients exhibited a more diverse microbiome with evenly distributed taxa. Their lesional microbiome profiles more closely resembled the contralateral skin microbiome and were enriched for taxa such as *Cutibacterium* (13% RAmean), *Bacillus* (19% RAmean), and *Kocuria* (13% RAmean). This microbiome cluster of lesions, M0, had a heterogeneous skin-like microbiota compared to the other clusters (Fig. 2E and 2F). Because host-level factors can influence the skin microbiota, we used multivariate linear regression analysis to identify statistically significant associations between the microbiome clusters and clinical metadata. These analyses did not detect any statistically significant associations between these parameters (Supplemental Fig. 2B). However, we cannot rule out the size of the cohort as a limiting factor in detection. Together, these data show that infection by *L. braziliensis* drives the lesional microbiota to a variety of configurations, in which *Staphylococcus* is the most frequent, consistent with our previous findings ^9^.

### Stratification by microbiome profile reveals signatures associated with delayed clinical resolution

We next investigated whether lesional microbiome clusters were associated with delayed clinical resolution. We designated the M0 cluster as a reference control, due to the heterogenous, skin-like microbiota characterizing these lesions. When we compared the healing times of clusters M1-M7 to the M0 cluster, we observed a trend toward delayed clinical resolution, P=0.09 (Fig. 3A top). When we stratified the samples by microbiome cluster, M6 lesions were delayed in healing time compared to M0, P=0.02 (Fig. 3A bottom). The other microbiome clusters did not significantly differ in clinical outcome compared with M0. However, these clusters contained fewer samples resulting in less statistical power (Supplemental Fig. 3A). Interestingly, lesions from the M6 and M7 clusters presented increased *L. braziliensis* parasite burdens, measured by qPCR, compared to lesions in the M0 cluster (Fig. 3B). This is consistent with our previous findings that increased parasite burdens are associated with treatment failure ^12^.

**Fig. 3.**
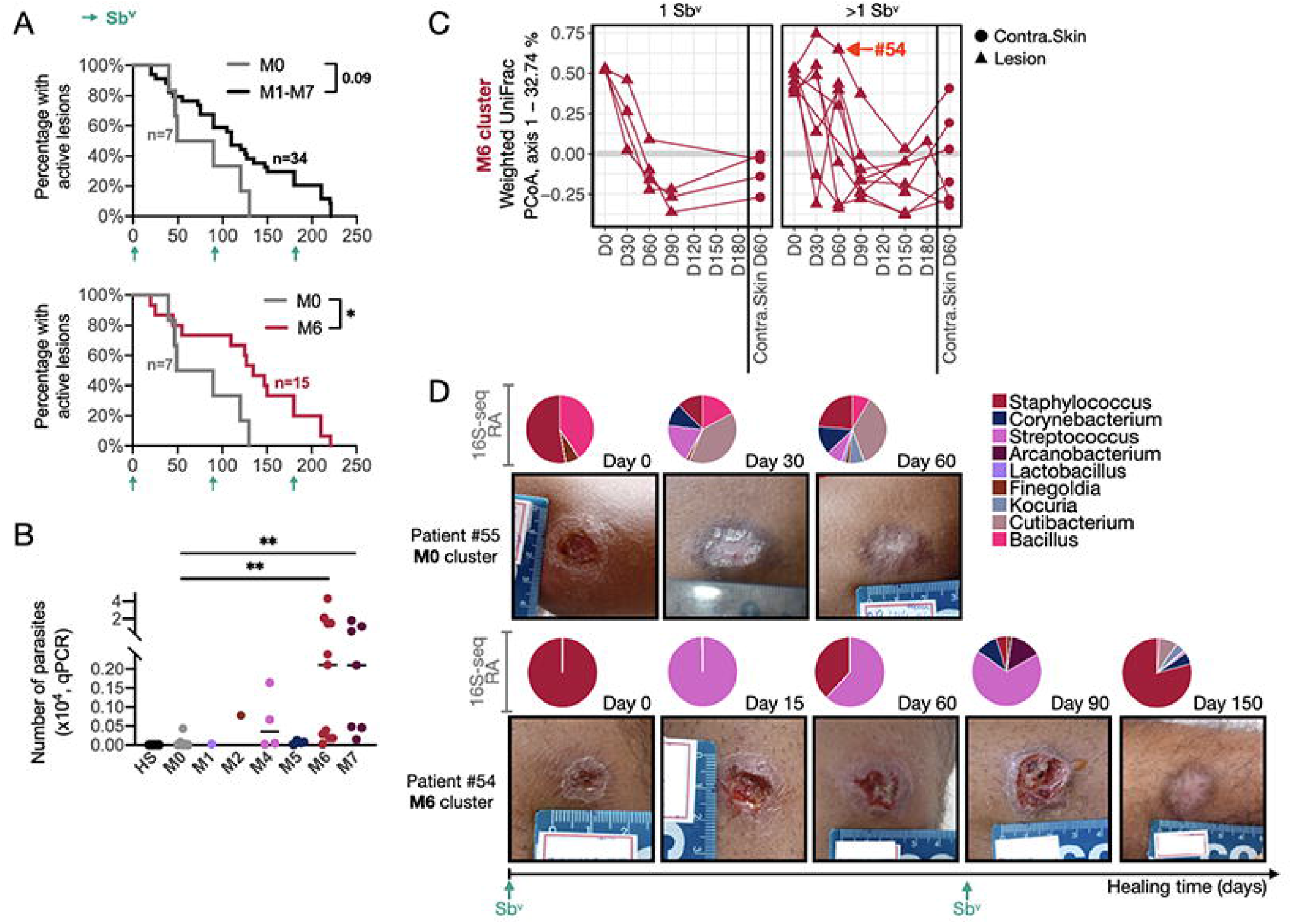
Predominant colonization with Staphylococcus is associated with delayed healing time and disturbed microbiome recovery. A) Survival curves for the healing time for lesions of patients from the M1-M7 clusters combined vs. M0 cluster, as well as separately M6 vs. M0 cluster. Patients who received an alternative form of treatment from an ongoing clinical trial in the clinic were not considered or added to the clinical outcome-related analysis. Log-rank (Mantel-Cox) test was used to calculate the statistical significance, *P=0.02. B) As measured by qPCR, the absolute number of parasites detected in 4-mm punch biopsies from CL lesions. T-test was used for statistical testing **P<0.01. C) PCoA showing component 1 calculated from weighted UniFrac dissimilarity analysis, including longitudinal swab samples from the M6 cluster (Day0-Day180) and swab samples from the contralateral skin of the same patient. Patients were divided for analysis according to the number of Sb^v^ rounds required for complete healing (1 vs. >1 round of Sb^v^). The connecting lines indicate samples collected from the same patient over time. D) Two representative examples of photographs taken from M0 (Patient #55) and M6 (Patient #54) lesion clusters. The relative abundance pie charts from 16S-seq profiling evaluated over time are indicated on top of the associated photos.

We next considered the succession of lesional microbial communities over time and the relationship with clinical course. We calculated weighted UniFrac dissimilarity metrics on swab samples collected at the initial visit to the clinic (before treatment) and at follow-up visits (Fig. 3C and Supplemental Fig. 3B). Focusing this analysis on the M6 cluster, we compared patients who required >1 round of Sb^v^ treatment to those who cured after a single treatment. In patients cured with just one round of Sb^v^, the lesional microbiome structure consistently shifted by day 60 to more closely resemble the contralateral skin community structure (Fig. 3C-left panel). Patients that required >1 round of Sb^v^ did not recover lesional signatures similar to skin communities by the end of follow-up (Fig. 3C-right panel). We also found that patients who required >1 round of Sb^v^ to heal exhibited individual dramatic microbiome structure shifts throughout the course of lesion development (Fig. 3C). A comparison of patients # 54 and # 55 provides a clear example of these dynamics. Patient #54, initially from the M6 cluster (*Staphylococcus* dominance), required 2 rounds of Sb^v^ treatment and did not heal until day 150. During this time, the lesional microbiome shifted to a *Streptococcus* predominant colonization (similar to the M4 cluster profile) but at the time of cure returned to 80% *Staphylococcus* relative abundance (Fig. 3D). In contrast, patient #55 from the M0 cluster required only one round of Sb^v^ and cured by day 60, and at the time of cure had a skin microbiome made up of taxa associated with a healthy microbiome, including *Cutibacterium, Bacillus* and *Kocuria* (Fig. 3D). Notably, weighted UniFrac dissimilarity analysis calculated with the longitudinal data from the contralateral skin of M6 patients revealed that the anti-parasitic Sb^v^ did not have a significant effect on the skin microbiome (Supplemental Fig. 3C). Thus, the longitudinal changes we observe in the lesional microbiome are unlikely driven by therapy alone.

### Altered microbiomes are associated with enhanced pro-inflammatory lesional gene expression

We hypothesized that different configurations of the lesional microbiome would drive distinct inflammatory and immune responses. To test this, we performed a combination of principal component analysis (PCA) (Supplemental Fig. 4) and DGE analysis (Supplemental Table 5) to examine the relationship between microbiome clusters and lesional gene expression (Fig. 4). We first compared M4, M5, M6 and M7 clusters with the M0 reference cluster to identify DEGs (separate analysis), combined the DEGs results and performed hierarchical clustering to identify gene modules. DGE analyses were not performed with lesions from M1-M3 clusters due to small sample sizes in these clusters; however, they were included for unsupervised HC classification. The dendrogram structure of the dendrogram suggested 2 major gene modules, Module 1 with genes overexpressed in the M0 cluster, and Module 2 with genes overexpressed in the M4-M7 clusters. GO analysis revealed that Module 1 was significantly enriched for *cell adhesion, Wnt signaling* and *nervous system development* gene signatures (Fig. 4B top and Supplemental Table 6), suggesting the induction of wound healing and skin repair processes. Module 2 was significantly enriched for innate pro-inflammatory signatures, such as *granulocyte/myeloid/lymphocyte-associated chemotaxis, response to cytokines such as IFN-y, response to LPS*, and *apoptotic processes* (Fig. 4B bottom and Supplemental Table 7). Examining the top 10 DEGs revealed that pro-inflammatory-related genes, including *IL1A* and *IL1B*, were overrepresented in the lesional transcriptional profiles of M1-M7 compared to M0 lesions (Fig. 4C). These findings suggest that deviations in the lesional microbiome away from the skin-like M0 profile are associated with enhanced pro-inflammatory gene expression.

**Fig. 4.**
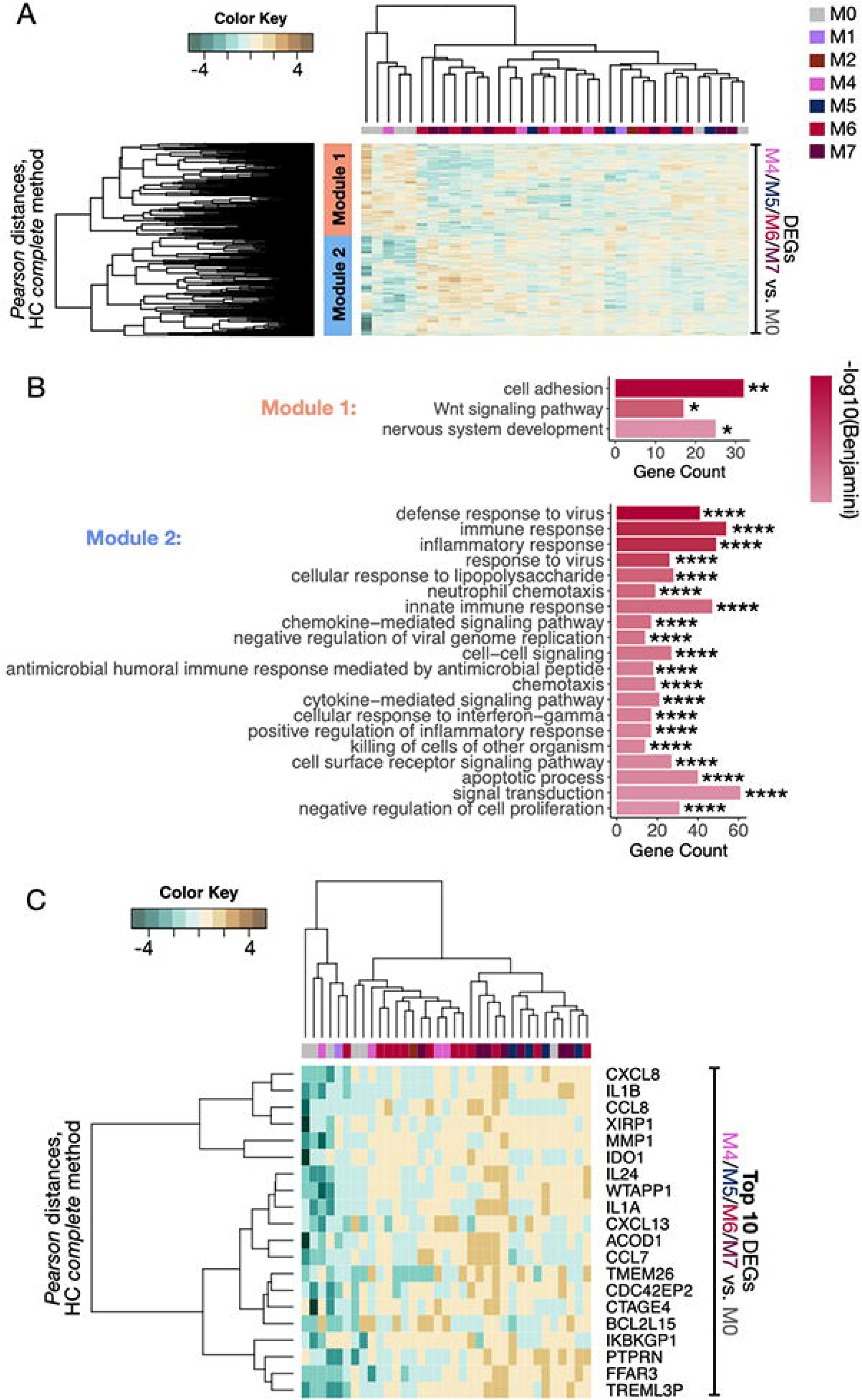
An altered microbiome is associated with enhanced pro-inflammatory gene expression. A) Separate DGE analyses were performed between lesions from the reference cluster M0 and clusters M4, M5, M6, and M7. The combined DEGs were then used to classify all lesions according to their transcriptional profiles by unsupervised HC (P<0.05 and FC>1.5). DGE analyses were not performed with M1-M3 due to insufficient sample size; however, they were included for unsupervised HC classification. Two gene modules were captured by Pearson’s distances (1-orange and 2-blue). B) GO analysis for Biological Processes (BP) terms was carried out in the two gene-modules from A. Bar graphs represent the number of genes identified in the GO genesets, and the color scale represents the statistical strength calculated by Benjamini-Bonferroni multiple correction testing. *P<0.05, **P<0.01, ****P<0.0001. C) Top 10 genes from each DGE analysis performed between the lesions from clusters M4, M5, M6, and M7 compared to M0 were concatenated and used to classify all the CL lesions according to their transcriptional profiles by HC. GO, Gene Ontology; DGE, Differential gene expression. DEGs, differentially expressed genes; HC, Hierarchical clustering.

### The magnitude of host pro-inflammatory responses correlates to the type of lesional microbiota

Because we observed a pro-inflammatory gene expression signature across all distinct microbiome clusters, we next asked if the magnitude and type of pro-inflammatory transcriptional programs might differ according to microbiome cluster. To test this, we used a previously described dataset-reductive computational workflow ^12^ (Fig. 5A). Briefly, to enhance statistical power, we focused on genes whose expression was highly variable between the lesions and were therefore termed variable transcripts associated with lesions (ViTALs). We reduced the list of ViTALs to principal components (PCs) from PCA (Fig. 5A and Supplemental Table 8).

**Fig. 5.**
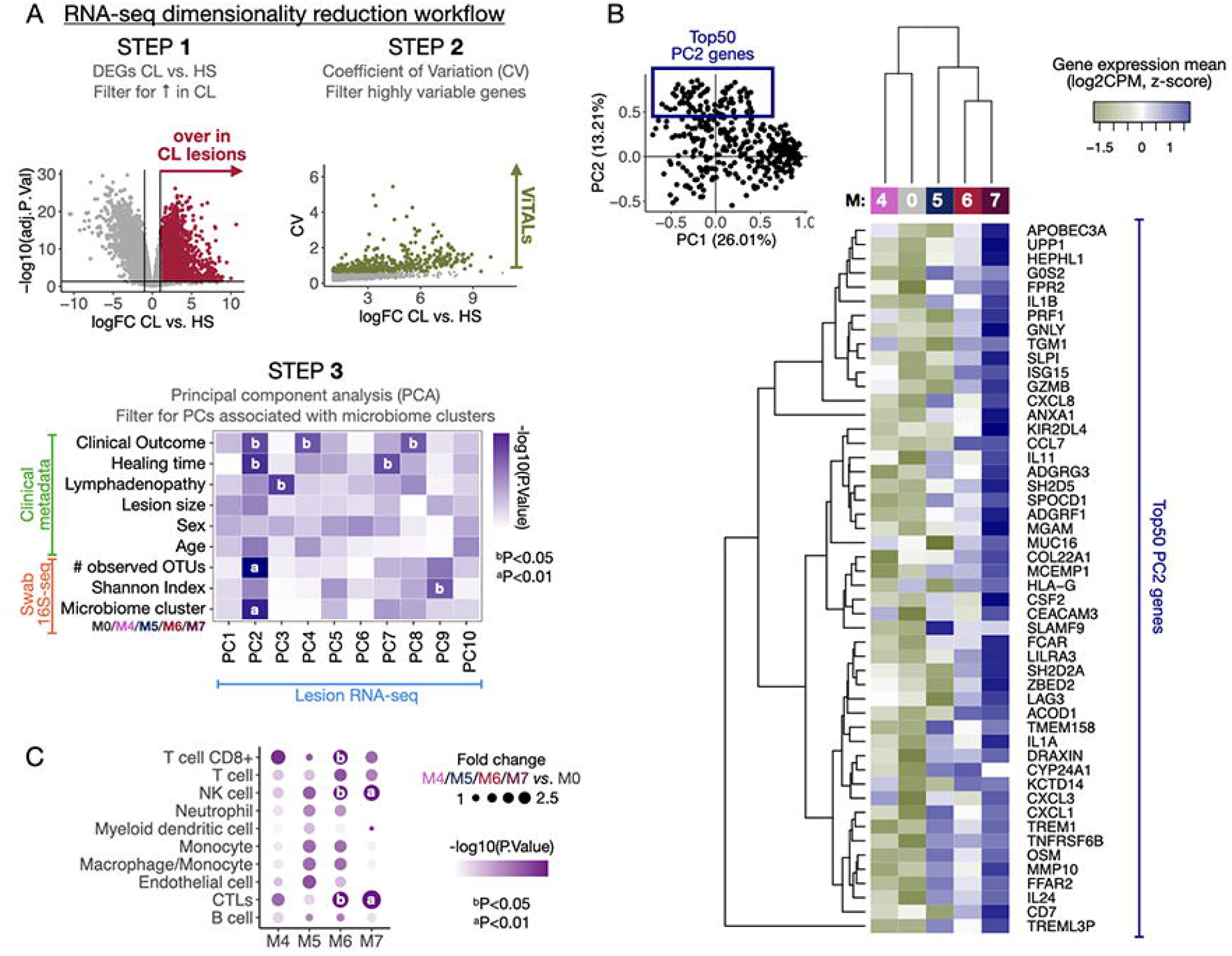
The distinct CL lesional microbiomes impact the pro-inflammatory gene expression profiles in different magnitudes. A) A dataset dimensionality reduction computational workflow was performed to integrate output parameters from the three primary datasets included in this study (RNA-seq, 16S-seq, and clinical metadata). Briefly, the gene expression matrix from the RNA-seq was reduced to a list of ∼400 genes overrepresented in CL vs. HS (step 1), displaying high variability among CL samples as calculated by the Coefficient of Variation, CV (step 2). Those genes are ViTALs and were further reduced to PCs in a PCA (step 3). A multivariate linear regression analysis was performed between the PCs and the 16S-seq parameters, and clinical metadata reduced output features. B) Top 50 PC2-explained genes are displayed in a heatmap and classified the transcriptional profiles by the microbiome clusters M4-M7 as calculated with HC. C) MCP-counter was used to estimate the cell population abundances from the RNA-seq dataset. A Differential cell-population analysis was performed, and the results from M4, M5, M6, and M7 vs. M0 are displayed as fold changes and P values. PCA, Principal Component Analysis. PC, Principal components. ViTALs, highly variable CL-associated transcripts. HC, hierarchical clustering.

These PCs were then associated with the 16S-seq and clinical metadata variables with multivariate linear regression analysis (Fig. 5A). PC2 had the strongest association with microbiome variables, including microbiome cluster and the number of observed OTUs, a measure of alpha diversity. PC2 was also associated with clinical outcomes (>1 round of Sb^v^) and increased healing times (Fig. 5A step 3, bottom). Though PC1 accounted for a greater amount of variation in the dataset (26.01%, compared to 13.21% for PC2), it was not associated with clinical metadata or microbiome variables. We thus focused on defining the genes that were most strongly associated with PC2.

The top 50 genes associated with PC2 included genes encoding for cytotoxicity-associated effector molecules and receptors such as PRF1, GNLY, GZMB, and KIR2DL4 (Fig. 5B). These genes, along with APOBEC3A, ISG15, and LILRA3, were previously identified as potential biomarkers for delayed treatment outcome ^12^. Other genes associated with PC2 encoded for neutrophil chemotaxis and effector functions, such as CEACAM3, CXCL1 and CXCL3; and genes encoding for pro-inflammatory cytokines such as IL-1α, IL-1β, OSM, IL-24 and SLPI (Fig. 5B). In most cases, these top 50 genes were most highly expressed in lesions from microbiome cluster M7 and were least expressed in lesions from microbiome cluster M0.

Though PC1 was not associated with clinical outcome or microbiome metrics, genes associated with PC1 included B cell response-related gene expression patterns (Immunoglobulins and B cell receptor-encoding genes) (Supplemental Fig. 5). Together, these findings support a relationship between the lesional microbiota, clinical outcome, and pro-inflammatory and cytotoxic transcriptional signatures, but not with B cell-related responses.

Because we observed highly variable pro-inflammatory lesional expression profiles across the microbiome clusters, we reasoned that immune cell populations might similarly differ. We used the Microenvironment Cell Populations-counter (MCP-counter) method, which allows quantification of the absolute abundance of immune cells in bulk RNA-seq samples on a gene marker basis. We observed increased frequencies of myeloid, granulocyte and lymphocyte cell populations in M4-M7 vs. M0 (FC>0). CTLs and NK cells were estimated to be significantly elevated in the lesions of M6 and M7 clusters compared to the M0 cluster (P<0.01 for M7 and P<=0.05 for M6) (Fig. 5C). Furthermore, CD8+ T cells were estimated to be significantly increased in M6 cluster (P<0.05) (Fig. 5C). Taken together, these observations strongly suggest that lesions in the M6 and M7 microbiome clusters express clinical outcome-related pro-inflammatory transcriptional signatures.

### Staphylococcus aureus in lesion biopsies is associated with pathogenic inflammation and delayed healing time

We found that lesions falling into the M6 and M7 microbiome clusters were characterized by enhanced pro-inflammatory signatures and increased relative abundances of *Staphylococcus*. The genus *Staphylococcus* encompasses a range of species that differentially impact host immune responses. For example, *S. epidermidis* is a critical skin commensal in mammals that mediates protective and adaptive immune responses required for tissue homeostasis. In contrast, *S. aureus* is the most common cause of skin and soft tissue infections and comprises ⅔ of the *Staphylococcus* isolates cultured from lesions of patients in this cohort Fig. 6A and Supplemental Table 8). However, resolving species and strains of *Staphylococcus* based on short amplicons of the 16S rRNA gene is challenging. Furthermore, while swab specimens are attractive for the collection of lesional and wound microbiota because they are generally non-invasive, they may not reflect deeper tissue bacterial burden and/or viable and replicating bacteria. Therefore, we focused our subsequent analysis on the pathogen *S. aureus*, with a refined RNA-seq pipeline to identify *S. aureus* transcripts in lesion biopsies.

**Fig. 6.**
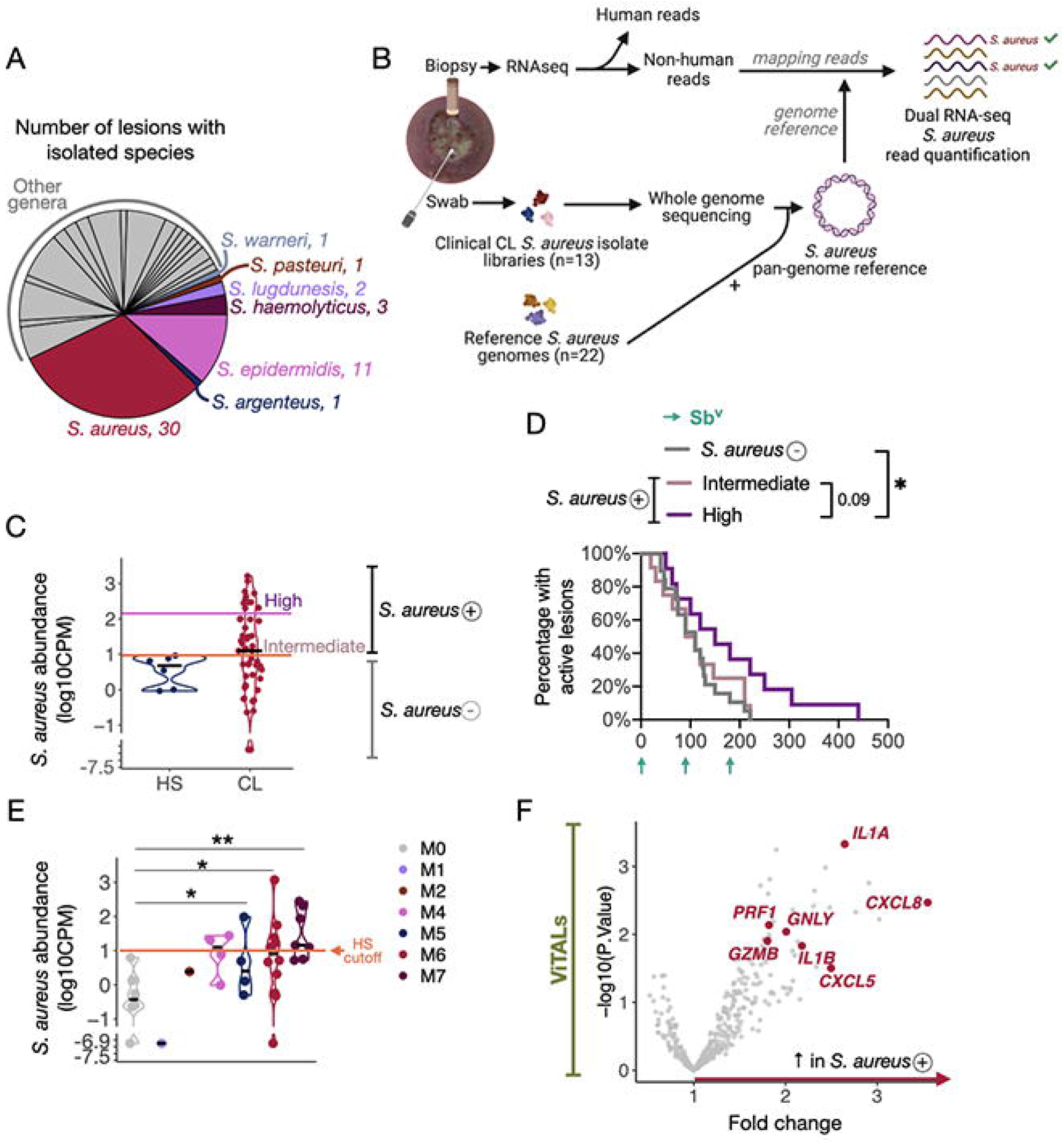
Dual RNA-seq analysis with an in-house S. aureus pan-genome reference identified the potential host pro-inflammatory mechanisms associated with S. aureus abundances. A) Name and number of clinical bacteria specimens isolated from 62 CL lesions. *Staphylococcus* spp. were annotated in the pie chart. B) In-house *S. aureus* pangenome-based dual RNA-seq workflow to quantify and estimate *S. aureus* abundances in CL lesions. The non-human reads from the 51 CL RNA-seq samples were mapped to our pan-genome reference, which is composed of genomes of 13 CL *S. aureus* clinical isolates and 22 *S. aureus* reference publicly available strains. C) Abundance levels quantified in the six healthy RNA-seq lesion samples served as threshold (log10CPM = 1) to divide CL lesions into the groups of increased *S. aureus* abundances *S. aureus* + lesions vs. *S. aureus* – lesions. 12 lesions presented overly high S. aureus abundances (FC = 18x) compared to 15 lesions with intermediate levels (threshold = 2 log10CPM). D) Survival curves for the healing time for *S. aureus* – lesions and high/intermediate *S. aureus* abundances (*S. aureus* + lesions). Patients who received an alternative form of treatment from an ongoing clinical trial in the clinic were not considered or added to the clinical outcome-related analysis. Log-rank (Mantel-Cox) test was used to calculate the statistical significance, *P<0.05. E) Abundance levels quantified in the CL RNA-seq lesion samples according to their corresponding 16S-Seq microbiome cluster classification. The pink horizontal line indicates the threshold based on HS samples (log10CPM = 1). T-test was used to calculate the statistical significance, *P<0.05 and **P<0.01. F) DGE analysis was performed with ViTALs between *S. aureus* + lesions vs. *S. aureus* – lesions (FC>1.5 and P<0.05). DGE, Differential gene expression analysis; GO, Gene ontology. FDR; False discovery rate.

To approach this, we first built a custom reference *S. aureus* pangenome derived from 13 *S. aureus* clinical isolates cultured from lesions in this cohort, improving our ability to detect *S. aureus* transcripts (Supplemental Fig. 6A, 6B). These isolates underwent whole genome sequencing (WGS) and together with 22 publicly available genomes from *S. aureus* reference strains, were used to construct the *S. aureus* pangenome (Fig. 6A and 6B and Supplemental Fig. 6A, 6B). Transcripts that failed to map to the human reference were then mapped to the *S. aureus* pangenome and quantified to estimate tissue burden. Relative to biopsies from uninfected, healthy skin, *S. aureus* transcripts were elevated in 27 lesions (Fig. 6C and Supplemental Fig. 6C). Moreover, 12 of those lesions expressed high *S. aureus* transcript abundances, around an 18x fold change compared to lesions with intermediate levels (Fig. 6C and Supplemental Fig. 6C). Importantly, we found that lesions with high abundances of *S. aureus* transcripts were delayed in healing when compared to lesions with abundances similar to intact skin (P<0.05) (Fig. 6D). Although the skin swabs did not show a dominance of *Staphylococcus* in the M7 cluster, *S. aureus* transcript abundances were increased in clinical isolates from these patients, suggesting that these bacteria may reside in deeper tissue not accessible by swabbing (Fig. 6E). Together, these findings indicate that the detection of *S. aureus* transcripts in lesional tissue is indicative of poor clinical outcome.

We next investigated the lesion’s host gene expression profiles that associated with transcriptionally active *S. aureus* in the lesion. We performed a DGE analysis on the previously curated list of ViTALS to compare *S. aureus*-positive vs. *S. aureu*s-negative lesions (Fig. 6F and Supplemental Table 9). Genes enriched in *S. aureus*-positive lesions included: *IL1A, IL1B, CXCL5, CXCL8, GNLY, PRF1* and *GZMB*, all of which we previously associated with more severe disease in mice and treatment failure in patients ^12^. Furthermore, these genes were similarly upregulated in lesions of the M7 and the M6 microbiome cluster, which were also characterized by high levels of *S. aureus* transcripts. These findings provide strong evidence for potential immune mechanisms in which *S. aureus* contributes to inflammation and clinical outcomes in leishmanial lesions.

### S. aureus mediates IL-1-dependent pathology in a murine model of L. braziliensis infection

Based on our findings in human lesions, we hypothesized that *S. aureus* potentiates *Leishmania*-infection-induced pro-inflammatory programs that can influence disease outcomes. To directly test this, we colonized B6 mice with *S. aureus* taken from a *L. braziliensis* patient (Supplemental Figure 6B). Mice were colonized with bacteria, and then infected with *L. braziliensis* parasites (Fig. 7A). *S. aureus* colonization led to an increase in skin thickness when compared to control mice (Fig. 7B). Correspondingly, we found that lesions from *S. aureus* colonized mice exhibited an increase in immune cells, particularly T cells and neutrophils (Fig. 7C, and Supplemental Figure 7). These results indicate that *S. aureus* colonization promotes increased disease in mice by promoting an excessive inflammatory response. Since we found that human lesion biopsies with increased levels of *S. aureus* had enhanced transcription of the genes encoding IL-1 family members, we next blocked IL-1β or IL-1 receptor to test if either were required for the increased pathology observed in mice. We found that neutralizing IL-1β or IL-1R significantly reduced the severity of the disease (Fig 7B, 7C). While the depletion of IL-1 signaling lessened pathology, there was a concomitant increase in the *S. aureus* burdens (Fig. 7D). This result indicates that the pathology was immune-mediated, rather than a direct effect of the bacteria. There was no significant change in the parasite burden in mice treated with anti-IL-1β or anti-IL-1R antibody (Fig. 7E). Together, these results implicate IL-1 as a critical factor driving increased disease severity in *S. aureus*-dominated lesions in mice and suggest that IL-1 can be a therapeutic target applicable to human cutaneous leishmaniasis.

**Fig 7.**
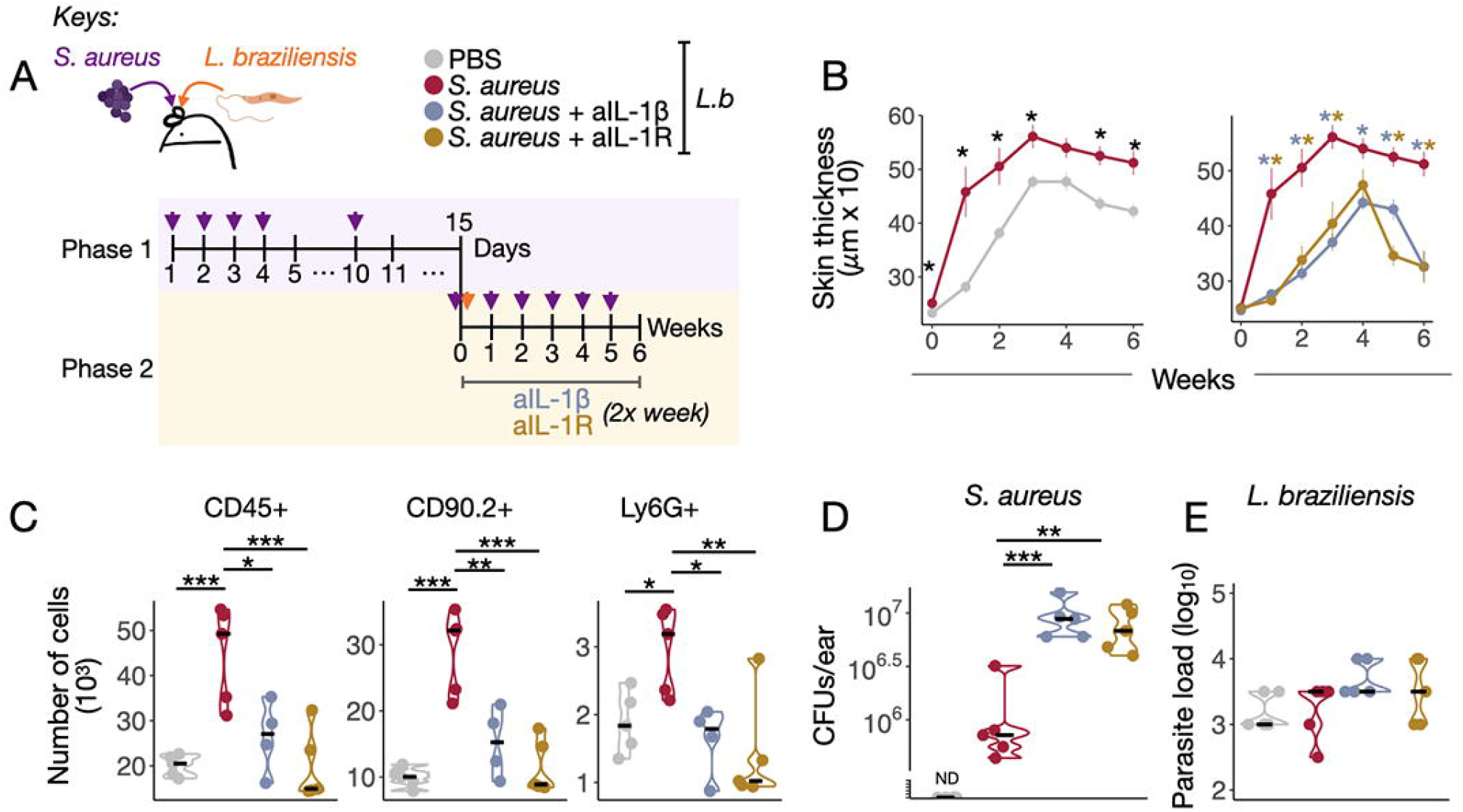
S. aureus and L. braziliensis co-infection increase the inflammatory responses in an IL-1 dependent manner. A) Schematic representation of *S. aureus* and *L. braziliensis* treatment protocol in C57BL/6 mice divided in Phase 1: pre-*S. aureus* colonization, and Phase 2: *L. braziliensis* infection course and blockade of IL-1β and IL-1R with monoclonal antibodies. B) Ear thickness measurements over time in mice. C) Number of CD45+, CD90.2+ and Ly6G+ cells recovered from the co-infected ears. D) Recovered pink *S. aureus* colony forming units (CFUs), and E) Parasite loads in the ear of different treatment groups in week six. Non-parametric Mann-Whitney test was used for statistical significance, *P<0.05, **P<0.01, ***P<0.001. Violin plots and median are represented in the plots when appropriate.

## DISCUSSION

*Leishmania braziliensis* infections are associated with chronic ulcerative lesions and poor responses to drug treatment targeting the parasite ^1–4,13^. Multiple factors could contribute to treatment failure ^13,14^, one of which is the upregulation of a cytolytic pathway mediated by CD8+ T cells and NK cells leading to inflammasome activation and IL-1β production ^12^. Here, we show that alterations in the skin microbiome also influence treatment outcomes. We analyzed a multi-omics dataset that included 62 patients infected with *L. braziliensis* to profile and associate the skin microbiome with host gene expression and clinical metadata. We found that most patients infected with *L. braziliensis* exhibited an altered skin microbiome in their lesions most often dominated with *Staphylococcus*, while in a smaller number of patients *Corynebacterium, Streptococcus*, or *Arcanobacterium* were dominant. Both the burden and the bacterial community structure influenced healing times, with *S. aureus* being an important species associated with delayed clinical outcomes. We also found that patients with higher levels of *S. aureus* exhibited increased expression of pro-inflammatory genes, including IL-1α and IL-1β. Finally, neutralizing IL-1β signaling in mice colonized with *S. aureus* resulted in reduced IL-1-dependent pathology following *L. braziliensis* infection. Taken together, these results suggest that the skin microbiome in leishmanial lesions influences the progression of the disease and may delay healing time due to increased levels of IL-1.

To evaluate the association between the skin microbiome and clinical outcome in cutaneous leishmaniasis patients we profiled the microbial composition of lesions and unaffected skin (contralateral skin from the same patient) by 16S-seq analysis. Most (84%) of the patients in this cohort developed alterations in their lesional microbiome. However, a few patients (16%) maintained a more diverse bacterial community similar to unaffected skin, which allowed us to compare the host transcriptional responses in these individuals with those observed in the lesions with altered microbiomes. Lesions with a modified microbiome were associated with seven distinct community structures (M1-M7), but the most common was characterized by predominant colonization with *Staphylococcus* (M6, 1/3 of patients in this cohort).

*Staphylococcus* was the genus most successfully isolated from lesions, as assessed by the number of clinical specimens isolated and cultured from the lesions. This observation suggests that *Staphylococcus* is particularly efficient in establishing itself in the ulcer caused by *L. braziliensis*, as previously reported in a separate cohort of *L. braziliensis*-infected patients by us ^9^ and others ^15–17^.

We used several computational approaches to investigate the interactions between the microbiome and host transcriptional profiles in cutaneous leishmaniasis. One involved a series of filtering steps to reduce the dimensionality of our datasets (16S-seq, RNA-seq, and clinical metadata) to integrate their main aspects (microbiome clusters and clinical outcome) and associate them with host gene expression by multivariate linear regression analysis. We also performed dual-RNA-seq analysis using an in-house *S. aureus* pangenome reference to estimate bacteria transcriptional abundances in each lesion. With these computational approaches, we identified a statistically significant correlation between the quantitative (total bacterial burden and *S. aureus* levels) and qualitative (specific microbiome community structures) factors with an increase in several pro-inflammatory genes. We identified that high expression of IL-1β gene is associated with bacterial alterations in leishmanial lesions, as well as a delay in response to therapy. Further, we demonstrated that IL-1β increased disease in mice infected with *S. aureus* and *L. braziliensis*. These findings indicate that *S. aureus* contributes to immunopathology in leishmaniasis in an IL-1-dependent manner. This role for IL-1 in promoting pathology is consistent with other studies in experimental murine leishmaniasis ^18–24^ and with our human studies where we found that *IL1B* levels in lesions from *L. braziliensis* patients are predictive of treatment failure ^12^. We previously identified two pathways that lead to IL-1β-dependent pathology, one of which is dependent upon blocking IL-10 and leads to decreased parasite numbers but increased IL-1β-dependent pathology ^24^. The other pathway is initiated by cytolytic T cells that promote cell death, leading to NLRP3 activation and subsequent release of IL-1β ^22,25^. Here we show that a dysregulated skin microbiome is an additional pathway leading to increased disease dependent upon IL-1β.

Biopsies from patients with high levels of *S. aureus* had elevated expression of cytolytic genes, such as those encoding *PRF1, GZMB*, and *GNLY*. Since increased *S. aureus* was associated with delayed healing, it is not surprising that cytolytic genes were also upregulated, as we previously found that their expression predicted treatment failure ^12^. Thus, while a cytolytic pathway in other contexts can promote the control of some types of bacteria ^26^, that did not seem to be the case in this cohort. Instead, the tissue damage mediated by cytolytic T cells may provide the environment for *S. aureus* to invade and replicate in this patient population. This would be consistent with the ability of *S. aureus* to colonize broken skin and soft tissue ^27,28^. However, the enrichment of genes associated with both cytolysis and IL-1 in *S. aureus*-dominant lesions is a unique feature in *L. braziliensis*-infected patients compared to other skin diseases complicated by *S. aureus* colonization and infection. In a similar analysis of atopic dermatitis (AD) and psoriasis skin, Fyrquist et al. found that AD skin exhibited high levels of *S. aureus*, accompanied by transcriptional activation of Th2-associated inflammatory pathways ^27^. As Th2 gene expression was not evident in this study, it appears that *S. aureus* can be associated with different inflammatory responses, depending on the context.

In addition to the *Staphylococcus* dominated profiles, less frequently observed configurations of the microbiota were also observed in some patients. For example, one microbiome cluster was characterized by *Arcanobacterium* genera (M7 cluster), and we successfully cultured and isolated *Arcanobacterium haemolyticum* from leishmanial lesions (Supplemental Table 5). Lesions from those patients exhibited aberrant pro-inflammatory gene expression relative to the whole cohort of biopsy samples, including cytolysis-related genes and IL-1α/β-encoding genes. Additionally, those lesions presented the highest abundance of *S. aureus* and parasites. *A. haemolyticum* is found in chronic wounds such as diabetic foot ulcers ^28^, as well as in soft-tissue infections from immunocompromised populations (carcinomas and other malignant tumors, rheumatism, hypertension, gout) ^28^ together with *S. aureus, Corynebacterium diphtheriae* and *Group A, C or G b-haemolytic streptococci* ^28,29^ *A. haemolyticum* produces phospholipase D, which protects it from *S. aureus*-related hemolysis, and co-infection can lead to cellular necrosis in HeLa cells ^30^. Little is known about the impact of this specific microbiota configuration of the microbiota on the immune responses associated with other skin infections. While the sample size of patients with *A. haemolyticum* was too low to associate with delayed healing, our results suggest that the *A. haemolyticum-S. aureus* community in leishmanial lesions may have a particularly detrimental impact on the clinical disease course.

Cutaneous leishmaniasis is an infectious disease characterized by complex interactions between the host and the parasite. Here, we demonstrate that cutaneous leishmaniasis leads to dramatic changes in the skin microbiome. Using an integrative approach employing clinical specimens and experimental murine infections, we directly demonstrate the microbiome’s critical role in the outcome of leishmanial infection. With resistance to Sb^v^ increasing in many endemic areas, the search for new treatment strategies is crucial. Here, we provide additional evidence supporting immunotherapy targeting IL-1 to reduce the inflammatory responses seen in cutaneous leishmaniasis. Further, our results provide a rationale for new therapies to influence the skin microbiome in patients, which might be accomplished by antibiotics, probiotics, and improved strategies for care of the leishmanial lesion.

## Supporting information

SupplementalTable1_LeishOmics_StudyDesign_medRxiv

SupplementalTable2_SaureusStrains_Pangenome

SupplementalTable3_DEGs_bacterial_burden

SupplementalTable4_GO_positive_bacterial_burden

SupplementalTable5_DEGs_microbiomeClusters

SupplementalTable6_GO_GeneModule1_MicrobiomeClusters

SupplementalTable7_GO_GeneModule2_MicrobiomeClusters

SupplementalTable8_IsolateLibrary

SupplementalTable9_DEGs_Saureus

## Data Availability

All data associated with this study are present in the paper or the Supplementary Materials. Raw sequence data are available at the Sequence Read Archive (SRA) as follows: bulk RNA-seq GEO BioProject PRJNA885131, 16S-seq and whole genome sequencing for S. aureus clinical isolates SRA BioProject PRJNA922957. Shell scripts and main R scripts are available in GitHub https://github.com/camilafarias112/Amorim_LeishMicrobiome. Tables from DGE analysis and GO analysis are available within this manuscript as Supplemental Tables.

## ACKNOWLEDGEMENTS

We profoundly thank the staff at Corte de Pedra, Salvador, Bahia, Brazil for assistance with patient screening and sample collection in this study. This work was funded by the following grants from the NIH (R01AI143790 to PS/EAG, R01NR015639 to EAG, and P50 AI030639 to E.M.C.). This research was also supported by the Penn Skin Biology and Disease Resource-based Center (Penn SBDRC supported by NIH/NIAMS P30AR069589). VML is supported by NIAMS F31AR079901 and JCH was supported by NIAMS F31AR079845 and the Penn Dermatology Research T32 Training Grant (NIH/NIAMS T32AR007465).

## AUTHORS CONTRIBUTIONS

C.F.A., D.P.B., F.O.N., E.A.G., and P.S. conceptualized the study; C.F.A., V.M.L., T.S., F.O.N., and J.C.H. participated in the methods and validated the results; C.F.A., V.M.L., T.S., and J.C.H. analyzed the results; C.F.A., V.M.L., T.S., F.O.N., J.C.H., D.P.B., P.S., and E.A.G. participated in the investigation; L.P.C., E.M.C., D.P.B., P.S., and E.A.G. provided resources and funding acquisition; C.F.A., V.M.L., T.S., and J.C.H. participated in data curation; C.F.A., D.P.B., P.S., and E.A.G. contributed to the writing of original draft, review and editing; C.F.A., D.P.B., P.S., and E.A.G. supervised this study.

## COMPETING INTERESTS

The authors declare that they have no competing interests.

## STAR METHODS

### Study design

Samples and metadata were collected from *L. braziliensis* patients at a Leishmaniasis Reference Center in Corte de Pedra, Bahia, Brazil. The diagnosis of leishmaniasis is made by the documentation of DNA of *L. braziliensis* by PCR, or isolation of parasites or by a typical lesion and one of the following: DTH to leishmania antigen and histopathology. Exclusion criteria included previous anti-leishmanial treatment, individuals under 18 years old, pregnancy, presence of other comorbidities, and a positive serologic test for human immunodeficiency virus (HIV). Informed consent was obtained from those enrolled in the study, after which lesions were measured and photographed. Lesions and contralateral skin were swabbed for 16S-seq microbiome profiling and to obtain bacterial isolates. Punch biopsies were collected from the border of the lesions and stored in RNAlater (Thermo Fisher Scientific). Additional biopsies from healthy controls were also collected. In most cases, different sample types were collected from the same patient. After collection of swabs and biopsies, patients were given standard-of-care treatment (daily intravenous injections of pentavalent antimony (Sb^v^); 20 mg/kg per day for 20 days). Patients were evaluated for lesion resolution on day 30 and every 30 days after treatment to assess healing. In a smaller cohort of patients swabs for 16S-seq microbiome profiling and lesion photographs were collected at days 30-210 Supplemental Fig. 1. This study was conducted according to the principles specified in the Declaration of Helsinki and under local ethical guidelines (Ethical Committee of the Faculdade de Medicina da Bahia, Universidade Federal da Bahia, Salvador, Bahia, Brazil, and the University of Pennsylvania Institutional Review Board).

### Swab processing and Microbiome profiling by 16S-Sequencing

Swab specimens from lesions and contralateral skin were collected in sterile 0.15 M NaCl with 0.1% Tween 20 in PBS 1x and stored at -20^°^C. Thirty-two negative control swab samples were exposed to air in the same room where the patients were seen and are called “environmental” samples. These samples were collected throughout patient recruitment and inclusion in this study. Genomic DNA extraction from swabs, library preparation, and 16S rRNA amplicon sequencing from swab specimens were all performed at the CHOP High-throughput Sequencing and Analytical Core of the PennCHOP Microbiome Program. DNA was extracted using the QIACube HT robotic workstation for nucleic acid purification (Qiagen) together with the DNeasy 96 PowerSoil Pro QIAcube HT kit (Qiagen), per standard protocol. Amplification of the 16S rRNA gene V1–V3 region was performed using primers FP 5′-AGAGTTTGATCCTGGCTCAG-3′ and RC 5-ATTACCGCGGCTGCTGG-3′, followed by library preparation, quantification, and pooling as previously described (PMID: 34437510). Sequencing was performed on the Illumina MiSeq using 300 bp paired-end chemistry. Extraction blanks and DNA-free water were subjected to the same amplification and purification procedure to assess potential environmental contamination. Samples with a total number of counts sequenced below the numbers detected in environmental control samples (<896 counts) were removed from further analysis accounting for 15 out of 374 total samples, most of them from the contralateral skin of patients. The QIIME2 pipeline was used to process and analyze 16S-sequencing data using qiime2 q2cli version 2020.8.0 ^31^. Briefly, samples were demultiplexed using demux and denoised using Dada2. Sequences were aligned using maaft, and phylogenetic trees were reconstructed using fasttree. Shannon Index alpha diversity metric was estimated using alpha-group-significance. Weighted UniFrac and Bray-Curtis beta diversity metrics were estimated using core-metrics-phylogenetic after samples were rarefied to 896 reads per sample, and p values were adjusted for multiple hypothesis testing using Benjamini-Hochberg false discovery rate (FDR) corrections. Taxonomy was assigned to sequences using q2-feature-classifier classify-sklearn against the Silva rRNA reference database, version 138 (silva-138-99-nb-classifier.qza) ^32^. Taxa were collapsed to the genus. OTUs not present in at least 5% of the entire dataset were filtered out. The microbiome clustering classification was performed in the top 10 taxa vs. lesion samples matrix in an untargeted manner, with hierarchical clustering and the Ward D2 agglomeration method.

### Biopsy processing and RNA-seq gene expression profiling

Lesion and healthy skin biopsies were homogenized with a MP tissue homogenizer (MP Biomedicals), and RNA was extracted using the RNeasy Plus Mini Kit (QIAGEN) according to the manufacturer’s instructions and used to prepare complementary DNA (cDNA) sequence-ready libraries using the Illumina TruSeq Total Transcriptome kit with Ribo-Zero Gold for eukaryotic cytoplasmic and mitochondrial rRNA depletion (Illumina). Quality assessment and quantification of RNA preparations and libraries were performed using an Agilent 4200 TapeStation and Qubit 3, respectively. Samples were sequenced five times to enhance sequencing depth on an Illumina NextSeq 500 to produce 75–base pair single-end reads with a total mean sequencing depth of 42 million reads per biopsy sample. Raw reads were mapped to the human reference transcriptome (Ensembl; Homo sapiens version 100) using Kallisto version 0.46.0, and MultiQC v.18 was used to check the quality of the alignment. All subsequent analyses were conducted using the statistical computing environment R version 4.1.0, RStudio version 1.4.1717, and Bioconductor version 3.13. Briefly, transcript quantification data were summarized to genes using the BiomaRt and tximport package and normalized using the trimmed mean of M values (TMM) method in edgeR. Genes with <1 CPM in at least six samples (the size of the smallest group of replicates, HS) were filtered out. Normalized filtered data were variance-stabilized using the voom function in limma, and Differential Gene Expression analysis was performed with linear modeling using limma after correcting for multiple testing using Benjamini-Hochberg FDR correction. DGE analyses were performed between microbiome clusters with >3 samples (M4-M7 vs. M0). GO analyses were carried out using DAVID Bioinformatics Resources (2021 Update) from NIAID/NIH and biological process terms and occasionally the Reactome, KEGG, and Biocarta Pathway Database, clustering genesets by annotation similarity. MCP-counter ^33^ and immundeconv ^34^ R packages were combined to estimate cell population abundances from the RNA-seq dataset. Survival curves with Log-rank (Mantel-Cox) testing were performed in GraphPad Prism version 8.

### Dimensional reduction workflow and integration of datasets

We reduced the dimensionality of three primary datasets included in the study, and the main output parameters were used for integrative multivariate linear regression analysis. The reductive workflows and output parameters are: 1) The biopsy whole transcriptome RNA-seq of around mapped human coding genes in a sample vs. gene matrix was reduced to a 401 gene list of “variable transcripts associated with lesions” (ViTALs) among lesion samples. This pipeline is described in our previous report ^12^ and available for reproducibility in the dockerized code “capsule” archived on Code Ocean (https://doi.org/10.24433/CO.5903311.v1); 2) The whole swab 16S-Seq dataset from samples collected at day 0 initiated at around 30.000 OTU matrix of patient samples post-QC-preprocessing and post-low abundance filtering step was collapsed to 86 individual, top frequent genera and reduced to top 10 abundant taxa in lesion samples, and finally to eight microbiome clusters output by unsupervised hierarchical clustering with Ward D agglomeration method (M0-M7). The number of observed OTUs and Shannon Indexes per swab sample were also included as microbiome parameters; 3) The clinical metadata parameters are: categorical (clinical outcome: 1 or >1 Sb^v^ rounds, sex, and presence of lymphadenopathy) and continuous variables (healing time in days, lesion size in mm^2^, and age in number of years). These output parameters were integrated using the *rexposome* framework ^35^. The rexposome outputs were modeled for visualization in R programming language using tidyverse R packages.

### Staphylococcus aureus isolate library, pan-genome reference, and quantification of S. aureus abundance in skin biopsies

Bacterial isolates were collected with a swab, stored immediately in cryotubes with freezing media (autoclaved, filtered Tryptic Soy Broth (TSB) dissolved in de-ionized water with 1% sterile Tween 80% and 15% glycerol) at -20^°^C. For bacterial identification, swabs were removed from TSB and streaked out onto blood agar plates (TSA with Sheep Blood Plate) (Remel, R01201) overnight at 37°C. Morphologically unique colonies were sub-cultured onto blood agar plates and individual bacterial species were identified using Matrix-assisted laser desorption/ionization-time of flight (MALDI-TOF) (Pennsylvania Animal Diagnostic Laboratory System, New Bolton Center). Following identification, *S. aureus* isolates were short-read whole-genome sequenced. *S. aureus* DNA was extracted and purified using the Quick-DNA Fungal/Bacterial Miniprep Kit (Zymo Research, D6005) and short-read paired-end Illumina Hiseq 2500 sequenced by the PennCHOP Microbiome Core. The In-house *S. aureus* pan-genome was built with the CL isolates and publicly available *S. aureus* reference genomes identifying the core and accessory genes with Roary ^36^ Supplemental Table 2. *S. aureus* transcript identification and quantification were performed in the CL lesion RNA-seq files by mapping the reads post-filtering out human reads using KneadData to the *S. aureus* pan-genome. *S. aureus* abundances per RNA-seq lesion sample represent the number of mapped counts per million (non-human) reads.

### Quantification of bacterial burden in the skin biopsies by qPCR

The bacterial burden was quantified by qPCR from the same cDNA libraries used for lesional RNA-seq transcriptional profiling. A standard curve was prepared, followed by a cDNA library from *S. aureus* (subsp. *aureus* Rosenbach, 502A, ATCC #27217). For this *S. aureus* standard curve, RNA was extracted by RNeasy Plus Mini Kit (QIAGEN), and cDNA conversion was prepared with High-Capacity RNA-to-cDNA Kit (Applied Biosciences). qPCR was carried out on a ViiA 7 machine (Applied Biosciences) using Power SYBR Green Master Mix (Applied Biosciences) and primers targeting the 16S ribosomal subunit (357F_534R; forward 5′-CTCCTACGGGAGGCAGCAG-3′ and reverse 5′-AGAGTTTGATCCTGGCTCAG-3′) ^37,38^. The qPCR results were normalized to the initial biopsy tissue weight. All reactions were carried out in duplicate, and data is represented as pg/mg biopsy.

### Quantification of parasite burden in the CL skin biopsies by qPCR

*L. braziliensis* burden in CL lesion biopsies was quantified as described previously ^12^.

### S. aureus colonization and Leishmania braziliensis infection

Mice were topically associated by applying up to 300 ml of suspended bacteria (10^8^-10^9^ CFUs) to the ears and back of the mouse using sterile cotton swabs, every day for a total of 4 days, once at day 10, once simultaneously with the *L. braziliensis* infection at day 15, and then once a week during the treatment protocol. For infection metacyclic promastigotes of *L. braziliensis* were isolated by Ficoll (Sigma) density gradient centrifugation and injected intradermally into the ear with 10^6^ *L. braziliensis* parasites. Some mice were injected with 500 μg anti-IL-β antibody or anti-IL-1R antibody (all purchased from BioXcell) twice a week for the duration of the experiment. Lesions were monitored by ear thickness. Parasite counts were performed by a limiting dilution assay.

### Tissue processing and flow cytometry analysis

To prepare single cell suspension, ventral and dorsal sheets of the ears were separated from the cartilage and incubated for 90 min in CO2 incubator at 37°C in 1 mL volume of RPMI 1640 (Invitrogen, Grand Island, NY, USA) containing 0.25 mg ml^-1^ Liberase TL (Roche Diagnostics, Chicago, IL, USA). The digested ears were passed through a 3 mL syringe to make single-cell suspension. The cells were filtered through 70 μm nylon mesh and washed in FACS buffer at 1500 rpm for 5 minutes. Cells were suspended in FACS buffer for further analysis. For surface staining the following antibodies were used at 1:100 dilutions in FACS buffer according to the manufacture’s specifications. CD45 (30-F11, eBiosciences), CD3 (17A2, eBiosciences), CD90.2 (53-2.1, eBiosciences), βTCR (H57-597, eBiosciences), CD4 (RM4-5, Biolegend), CD8 (YTS5167.7, eBiosciences), CD11b (M1/70, eBiosciences), Ly6G (1A8, eBiosciences), and Ly6C (AL-21, BD Pharmingen). For counting the cells AccuCount Fluorecent particles (Spherotech, Lake Forest, IL, USA) were used. The stained cells were run on BD FACSymphony(tm) *A3* (BD Biosciences, San Jose, CA, USA) and the acquired data were analyzed using FlowJo software (Tree Star, Ashland, OR, USA).

### Sequencing datasets and code availability

All data associated with this study are present in the paper or the Supplementary Materials. Raw sequence data are available at the Sequence Read Archive (SRA) as follows: bulk RNA-seq GEO BioProject PRJNA885131, 16S-seq and whole genome sequencing for *S. aureus* clinical isolates SRA BioProject PRJNA922957. Shell scripts and main R scripts are available in GitHub https://github.com/camilafarias112/Amorim_LeishMicrobiome. Tables from DGE analysis and GO analysis are available within this manuscript as Supplemental Tables.

## SUPPLEMENTAL FIGURE TITLES AND LEGENDS

*Supplemental Figure 1. Study design, clinical metadata, and dataset sample inventory*. A) A schematic summarizing the sample types and time points of collection for this study. B) Venn diagram indicates the number of biopsies and swabs for 16S-seq collected from the same lesion and patient. C) A heatmap describes the sample inventory for each participating patient and their associated clinical metadata.

*Supplemental Figure 2. Microbiome clusters definitions and association with clinical metadata*. A) Unsupervised hierarchical clustering (HC) analysis with Ward.D2 clustering method were calculated from a relative abundance matrix of top 10 taxa to classify samples into eight distinct microbiome profiles, or “clusters”. A consensus of relative abundance means per sample, and number of dominant taxa in the community (>80% of total dominance and >2, respectively) were used as thresholds to cluster the microbiome groups. B) Multivariate linear regression analysis was calculated between the Microbiome clusters (M0-M7) and clinical metadata using the *lm()* function in R.

*Supplemental Figure 3. Impact of different microbiome dysbiosis in the lesion healing time in patients infected by L. braziliensis*. A) Survival curves for the healing time for lesions of patients from M1, M2, M3, M4, M5, M7 vs. M0 cluster. Patients who received an alternative form of treatment from an ongoing clinical trial in the clinic were not considered or added to the clinical outcome-related analysis. Log-rank (Mantel-Cox) test was used to calculate the statistical significance. B) PCoA showing component 1 calculated from weighted UniFrac dissimilarity analysis, including longitudinal swab samples from the M1, M2, M3, M4, M5, M7 and M0 clusters (Day0-Day210) and swab samples from the contralateral skin of the same patient. Patients were divided for analysis according to the number of Sbv rounds required for complete healing (1 vs. >1 round of Sbv). The connecting lines indicate samples collected from the same patient over time. C) Antimony therapy does not affect unaffected skin microbiome (contralateral skin). PCoA calculated from weighted UniFrac dissimilarity analysis including longitudinal swab samples from the lesion and contralateral skin of CL patients at Day0 and Day90. Lines connect samples from the same patients. Only patients treated with Sb^v^ and with samples available from those two time points were included in this analysis.

*Supplemental Figure 4. Principal Component Analysis (PCA) on whole host transcriptome with patients with distinct microbiome clusters*. Principal Component Analysis (PCA) showing the principal component (PC) 1 and PC2 for RNA-seq data from CL lesions with microbiome cluster available, n=35 (characterized from the 16S-seq analysis) and Healthy skin (HS), n=7.

*Supplemental Figure 5. Principal Component Analysis (PCA) on ViTALs showing gene loadings*. PCA showing the PC1 and PC2 for ViTALs from CL lesions with microbiome cluster available, n=35 (characterized from the 16S-seq analysis). This PCA is part of the Step 3 from our dataset dimensionality reduction computational workflow. In pink, genes named as “Immunoglobulin”-encoding genes. PCA, Principal Component Analysis. PC, Principal components. ViTALs, highly variable CL-associated transcripts.

*Supplemental Figure 6. Staphylococcus isolates from CL lesions and associated clinical metadata and RNA-seq/16S-seq results*. A) 13 *S. aureus* isolates were short-read whole genome sequenced and combined with 22 publicly available *S. aureus* genomes to build out in-house *S. aureus* pangenome. A Hierarchical clustering was performed between the *S. aureus* genomes, together with a publicly available *S. epidermidis* genome to serve as comparison. B) Clinical metadata, RNA-seq and 16S-seq-associated data from the lesions where the CL S. aureus isolates originated from. C) Distribution of lesion biopsies according to the S. aureus transcript abundances (in log10 counts per million, CPM).

*Supplemental Figure 7. Number of CD90*.*2-, αβT cells and CD4+ T cells in S. aureus and L. braziliensis treatment protocol in C57BL/6 mice, with blockade of IL-1α, IL-1β and IL-1R with monoclonal antibodies*. Number of CD90.2-, αβT cells and CD4+ T cells recovered from the co-infected ears in week six. Non-parametric Mann-Whitney test was used for statistical significance, *P<0.05, **P<0.01, ***P<0.001. Violin plots and median are represented in the plots when appropriate.

## SUPPLEMENTAL TABLE TITLES AND LEGENDS

*Supplemental Table 1. Study design and clinical metadata for the cutaneous leishmaniasis patients included in this study*. Age in years, size of the lesion and delayed type-hypersensitive results in mm^2^. Treatment outcome: cure = 1 round of antimony, and failure = 2-3 rounds of antimony. Healing time represents the number of days to complete formation of closed scar since the start of the therapy. Microbiome cluster represents the assigned cluster based on 16S-seq analysis. Bacterial load as measured in the skin lesion biopsies by qPCR. *S. aureus* transcript abundances as measured by dual RNA-seq. *Leishmania braziliensis* load as measured in the skin lesion biopsies by qPCR.

*Supplemental Table 2. List of cutaneous leishmaniasis and publicly available Staphylococcus aureus strains included in the in-house S. aureus reference core genome*.

*Supplemental Table 3. Differential gene expression (DGE) analysis was performed with the continuous variable “bacterial burden” measured by qPCR from CL lesions*. DEG analysis was carried out by linear modeling using limma after correcting for multiple testing using Benjamini-Hochberg FDR correction testing.

*Supplemental Table 4. Gene Ontology (GO) annotation genesets for the 148 genes correlated positively with bacterial burden*. GO analyses were carried out using DAVID Bioinformatics Resources (2021 Update) from NIAID/NIH and biological process terms.

*Supplemental Table 5. Differential gene expression (DGE) analysis was performed between patients in the microbiome clusters M4, M5, M6, M7 vs. M0*. DEG analysis was carried out by linear modeling using limma, intercepting the M4/M5/M6/M7 cluster to M0. Correction for multiple testing was performed using Benjamini-Hochberg FDR testing.

*Supplemental Table 6. Gene Ontology (GO) annotation genesets for the genes included in Gene-Module 1*. GO analyses were carried out using DAVID Bioinformatics Resources (2021 Update) from NIAID/NIH and biological process terms.

*Supplemental Table 7. Gene Ontology (GO) annotation genesets for the genes included in Gene-Module 2*. GO analyses were carried out using DAVID Bioinformatics Resources (2021 Update) from NIAID/NIH and biological process terms.

*Supplemental Table 8. List of all bacterial species/strains isolated from this cohort of cutaneous leishmaniasis patients*.

*Supplemental Table 9. Differential gene expression (DGE) analysis was performed in the list of ViTALs, between S. aureus + and S. aureus – CL lesions*. DEG analysis was carried out by linear modeling using limma after correcting for multiple testing using Benjamini-Hochberg FDR correction testing.

## Notes

### Competing Interest Statement

The authors have declared no competing interest.

### Author Declarations

This study was conducted according to the principles specified in the Declaration of Helsinki and under local ethical guidelines (Ethical Committee of the Faculdade de Medicina da Bahia, Universidade Federal da Bahia, Salvador, Bahia, Brazil, and the University of Pennsylvania Institutional Review Board).

### Summary of Updates

The title was updated; The data availability paragraph was updated with new URLs;

