## SupplementalFigures for "Multi-omic profiling of cutaneous leishmaniasis infections reveals microbiota-driven mechanisms underlying disease severity"

*Supplemental Figure 1. Study design, clinical metadata, and dataset sample inventory*. A) A schematic summarizing the sample types and time points of collection for this study. B) Venn diagram indicates the number of biopsies and swabs for 16S-seq collected from the same lesion and patient. C) A heatmap describes the sample inventory for each participating patient and their associated clinical metadata.

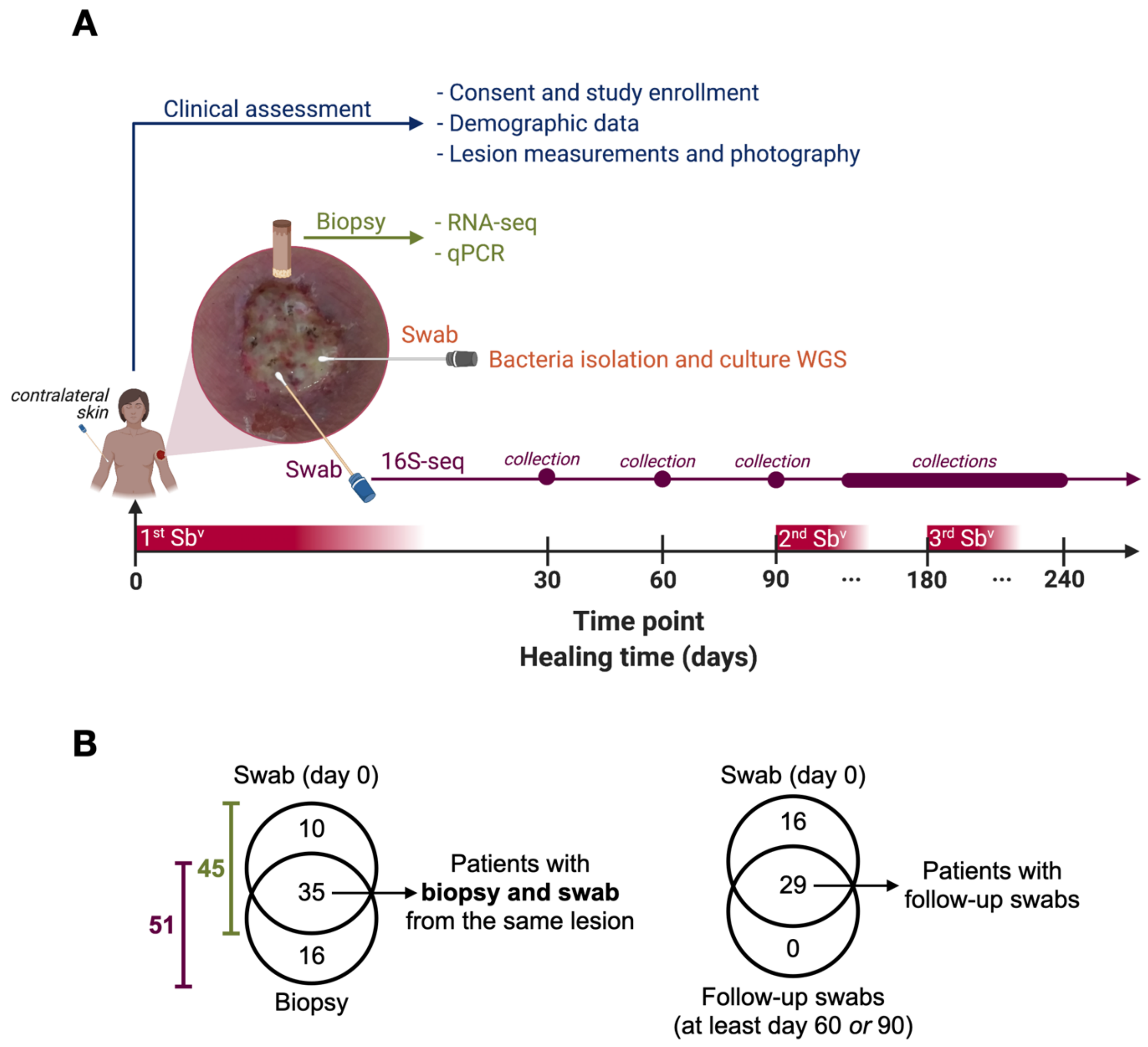

**C**

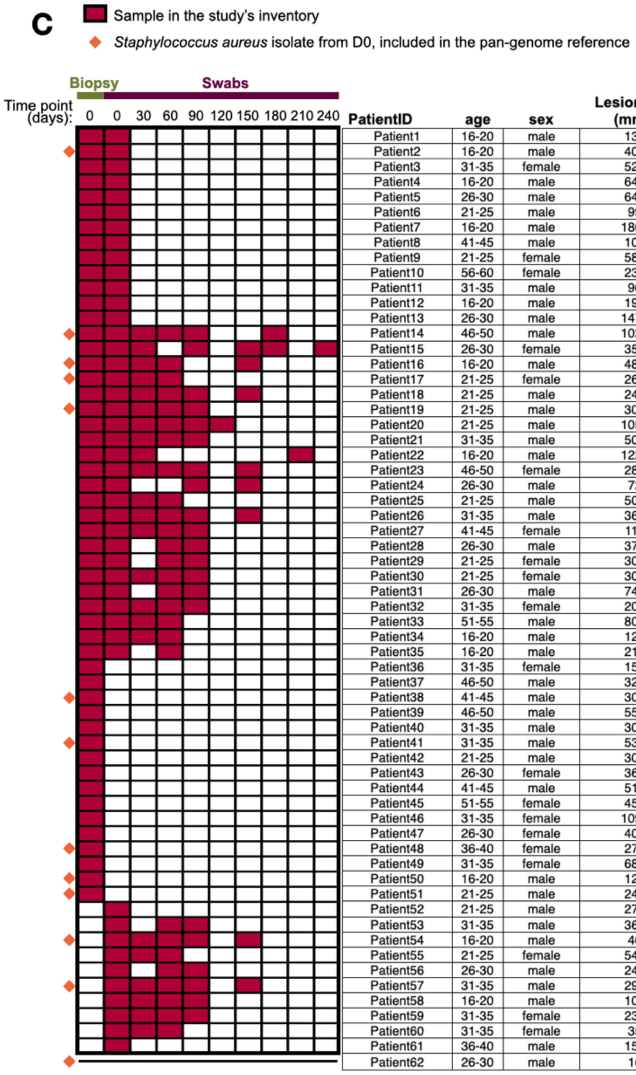


| PatientID | age | sex | Lesion size ($mm^2$) | Lymphadenopathy | DTH ($mm^2$) | Treatment Outcome*/** | Healing time (Days) | Treatment |
|---|---|---|---|---|---|---|---|---|
| Patient1 | 16-20 | male | 132 | yes | 132 | DOT | DOT | DOT |
| Patient2 | 16-20 | male | 400 | yes | 144 | Failure | 180 | antimony |
| Patient3 | 31-35 | female | 525 | no | 110 | Cure | 90 | antimony |
| Patient4 | 16-20 | male | 645 | yes | 120 | Failure | 210 | antimony |
| Patient5 | 26-30 | male | 646 | yes | 225 | DOT | DOT | DOT |
| Patient6 | 21-25 | male | 99 | yes | 140 | Cure | 49 | antimony |
| Patient7 | 16-20 | male | 1800 | no | 130 | Failure | 221 | antimony |
| Patient8 | 41-45 | male | 100 | yes | 130 | Failure | 147 | antimony |
| Patient9 | 21-25 | female | 580 | yes | 210 | Failure | 221 | antimony |
| Patient10 | 56-60 | female | 234 | yes | 144 | Cure | 98 | antimony |
| Patient11 | 31-35 | male | 90 | yes | 144 | DOT | DOT | DOT |
| Patient12 | 16-20 | male | 196 | yes | 120 | DOT | DOT | DOT |
| Patient13 | 26-30 | male | 1470 | yes | 225 | Cure | 90 | antimony |
| Patient14 | 46-50 | male | 1025 | yes | 120 | Failure | 221 | antimony |
| Patient15 | 26-30 | female | 357 | yes | 195 | DOT | DOT | DOT |
| Patient16 | 16-20 | male | 480 | yes | 130 | Failure | 120 | antimony |
| Patient17 | 21-25 | female | 264 | no | 252 | Cure | 45 | antimony |
| Patient18 | 21-25 | male | 240 | yes | 110 | Failure | 210 | antimony |
| Patient19 | 21-25 | male | 300 | yes | 225 | Cure | 72 | antimony |
| Patient20 | 21-25 | male | 1050 | yes | 182 | Failure | 110 | antimony |
| Patient21 | 31-35 | male | 500 | yes | 240 | Cure | 75 | antimony |
| Patient22 | 16-20 | male | 1221 | yes | 140 | Failure | 210 | antimony |
| Patient23 | 46-50 | female | 280 | yes | 210 | Failure | 127 | antimony |
| Patient24 | 26-30 | male | 72 | yes | 100 | Failure | 150 | antimony |
| Patient25 | 21-25 | male | 500 | yes | 121 | Cure | 110 | antimony |
| Patient26 | 31-35 | male | 360 | no | 144 | Failure | 125 | antimony |
| Patient27 | 41-45 | female | 110 | no | 165 | Cure | 75 | antimony |
| Patient28 | 26-30 | male | 375 | yes | 99 | Failure | 110 | antimony |
| Patient29 | 21-25 | female | 306 | yes | 132 | Cure | 90 | antimony |
| Patient30 | 21-25 | female | 300 | no | 225 | Cure | 20 | antimony |
| Patient31 | 26-30 | male | 740 | no | 90 | Cure | 47 | antimony |
| Patient32 | 31-35 | female | 200 | yes | 240 | Failure | 130 | antimony |
| Patient33 | 51-55 | male | 800 | yes | 323 | Cure | 40 | antimony |
| Patient34 | 16-20 | male | 120 | no | 832 | Cure | 40 | antimony |
| Patient35 | 16-20 | male | 216 | yes | 195 | Failure | 120 | antimony |
| Patient36 | 31-35 | female | 150 | no | 180 | DOT | DOT | DOT |
| Patient37 | 46-50 | male | 323 | no | 225 | DOT | DOT | DOT |
| Patient38 | 41-45 | male | 300 | yes | 192 | Cure | 50 | antimony |
| Patient39 | 46-50 | male | 550 | yes | 169 | DOT | DOT | DOT |
| Patient40 | 31-35 | male | 300 | no | 120 | Failure | 305 | antimony |
| Patient41 | 31-35 | male | 532 | no | 99 | Failure | 120 | antimony |
| Patient42 | 21-25 | male | 300 | yes | 121 | Cure | 30 | antimony |
| Patient43 | 26-30 | female | 360 | yes | 156 | DOT | DOT | DOT |
| Patient44 | 41-45 | male | 513 | yes | 100 | Failure | 180 | antimony |
| Patient45 | 51-55 | female | 450 | no | 225 | Cure | 70 | antimony |
| Patient46 | 31-35 | female | 1092 | no | 120 | Failure | 440 | antimony |
| Patient47 | 26-30 | female | 405 | no | 90 | Failure | 250 | antimony |
| Patient48 | 36-40 | female | 270 | no | 156 | Failure | 100 | antimony |
| Patient49 | 31-35 | female | 684 | no | 156 | Failure | 150 | antimony |
| Patient50 | 16-20 | male | 120 | yes | 81 | Cure | 63 | antimony |
| Patient51 | 21-25 | male | 240 | yes | 204 | Cure | 64 | antimony |
| Patient52 | 21-25 | male | 275 | yes | 285 | Failure | 220 | antimony |
| Patient53 | 31-35 | male | 360 | yes | 196 | Cure | 55 | antimony |
| Patient54 | 16-20 | male | 40 | yes | 56 | Failure | 135 | antimony |
| Patient55 | 21-25 | female | 540 | yes | 225 | Cure | 40 | antimony |
| Patient56 | 26-30 | male | 240 | yes | 195 | Cure | 37 | antimony |
| Patient57 | 31-35 | male | 294 | yes | 225 | Failure | 180 | antimony |
| Patient58 | 16-20 | male | 108 | yes | 156 | Cure | 25 | antimony |
| Patient59 | 31-35 | female | 230 | no | 180 | Failure | 180 | antimony |
| Patient60 | 31-35 | female | 35 | yes | 240 | Cure | 20 | antimony |
| Patient61 | 36-40 | male | 154 | no | 100 | Failure | 105 | antimony |
| Patient62 | 26-30 | male | 16 | yes | 272 | Failure | 130 | antimony |

*DOT = patients were treated with an alternative ongoing Drug On Trial at the endemic area

**Cure = 1 round of antimony, Failure = 2-3 rounds of antimony

**Table also as Supplemental Table 1**

*Supplemental Figure 2. Microbiome clusters definitions and association with clinical metadata.* A) Unsupervised hierarchical clustering (HC) analysis with Ward.D2 clustering method were calculated from a relative abundance matrix of top 10 taxa to classify samples into eight distinct microbiome profiles, or "clusters". A consensus of relative abundance means per sample, and number of dominant taxa in the community (>80% of total dominance and >2, respectively) were used as thresholds to cluster the microbiome groups. B) Multivariate linear regression analysis was calculated between the Microbiome clusters (M0-M7) and clinical metadata using the *lm()* function in R.

**A**

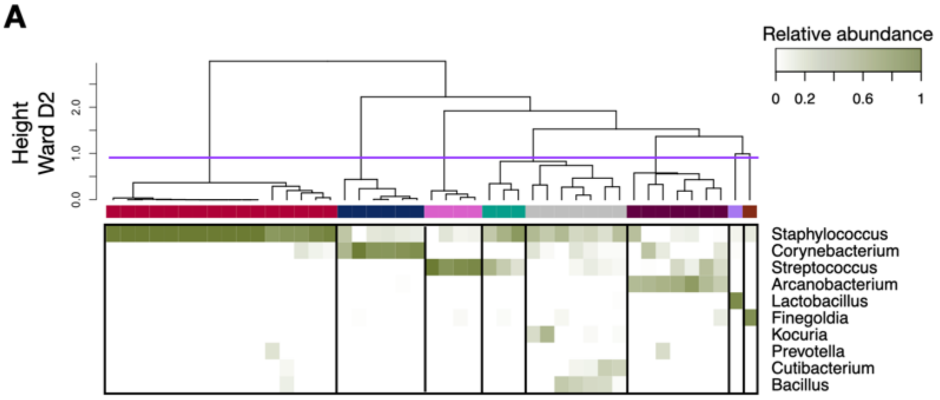


**B**

**Multivariate linear regression model:**
**Microbiome cluster vs. Age, Sex, Lesion size, DTH, Lymphadenopathy**

| | Estimate | Std.Error | t value | Pr(>\|t\|) |
|---|---|---|---|---|
| (Intercept) | 4.014651 | 2.5935503 | 1.548 | 0.13 |
| Age | 0.0010747 | 0.0395538 | 0.027 | 0.978 |
| Sex | 0.6015822 | 0.8833021 | 0.681 | 0.5 |
| Lesion size (mm2) | 0.0006427 | 0.0010042 | 0.64 | 0.526 |
| DTH (mm2) | -0.0015454 | 0.0033215 | -0.465 | 0.644 |
| Lymphadenopathy | -0.4494788 | 0.9347437 | -0.481 | 0.633 |

Microbiome cluster residuals: Min=-4.6844; 1Q=-1.3933; Median=0.9822; 3Q=1.6368; Max=2.8452
Residual standard error: 2.466 on 39 degrees of freedom
Multiple R-squared=0.0483; Adjusted R-squared=-0.07371;
F-statistic=0.3959 on 5 and 39 DF; p-value: 0.8486

*Supplemental Figure 3. Impact of different microbiome dysbiosis in the lesion healing time in patients infected by L. braziliensis.* A) Survival curves for the healing time for lesions of patients from M1, M2, M3, M4, M5, M7 vs. M0 cluster. Patients who received an alternative form of treatment from an ongoing clinical trial in the clinic were not considered or added to the clinical outcome-related analysis. Log-rank (Mantel-Cox) test was used to calculate the statistical significance. B) PCoA showing component 1 calculated from weighted UniFrac dissimilarity analysis, including longitudinal swab samples from the M1, M2, M3, M4, M5, M7 and M0 clusters (Day0-Day210) and swab samples from the contralateral skin of the same patient. Patients were divided for analysis according to the number of Sbv rounds required for complete healing (1 vs. >1 round of Sbv). The connecting lines indicate samples collected from the same patient over time. C) Antimony therapy does not affect unaffected skin microbiome (contralateral skin). PCoA calculated from weighted UniFrac dissimilarity analysis including longitudinal swab samples from the lesion and contralateral skin of CL patients at Day0 and Day90. Lines connect samples from the same patients. Only patients treated with $Sb^v$ and with samples available from those two time points were included in this analysis.

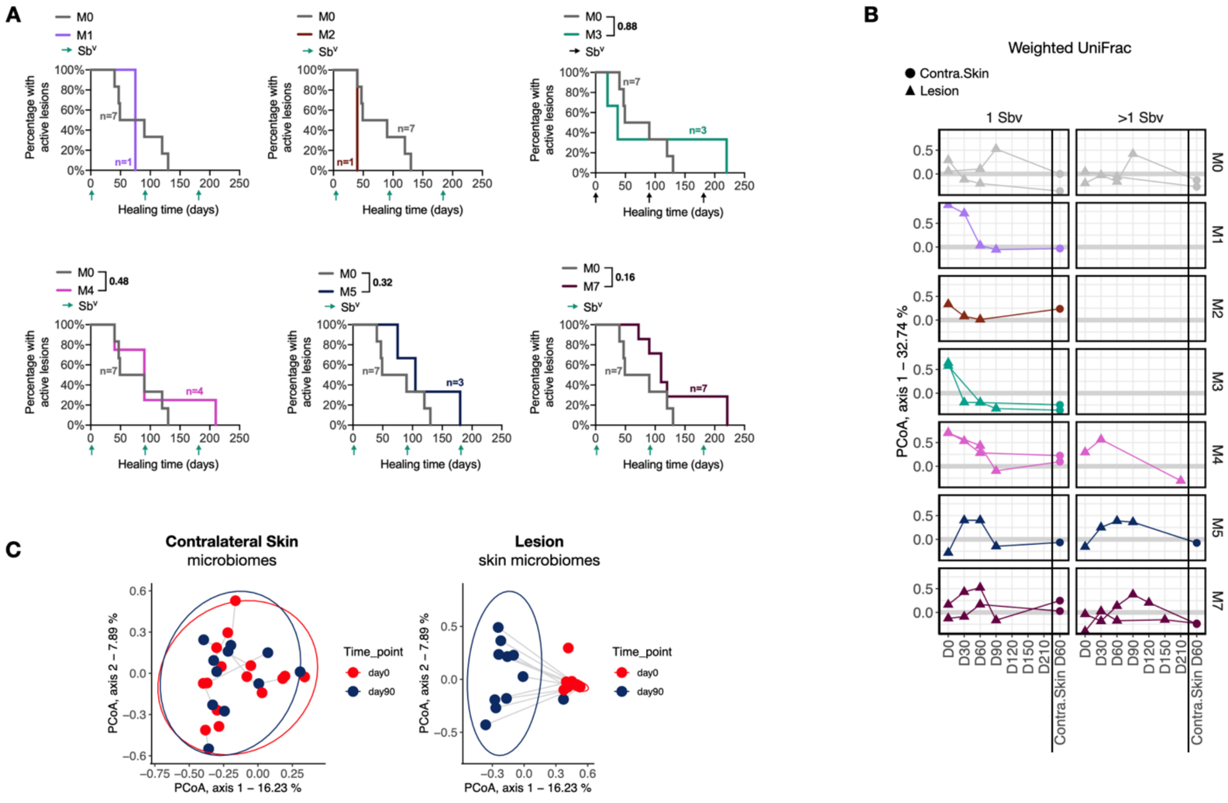

*Supplemental Figure 4. Principal Component Analysis (PCA) on whole host transcriptome with patients with distinct microbiome clusters.* Principal Component Analysis (PCA) showing the principal component (PC) 1 and PC2 for RNA-seq data from CL lesions with microbiome cluster available, n=35 (characterized from the 16S-seq analysis) and Healthy skin (HS), n=7.

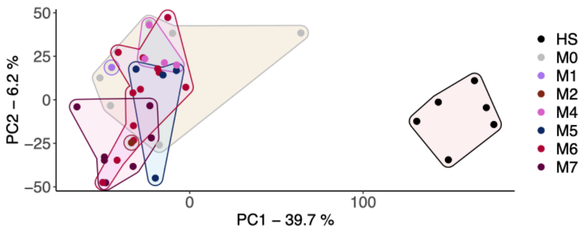


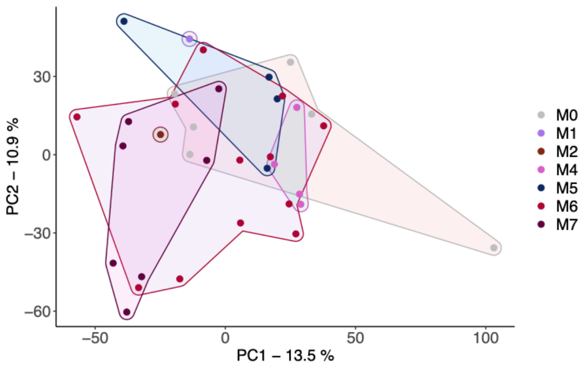

*Supplemental Figure 5. Principal Component Analysis (PCA) on ViTALs showing gene loadings.* PCA showing the PC1 and PC2 for ViTALs from CL lesions with microbiome cluster available, n=35 (characterized from the 16S-seq analysis). This PCA is part of the Step 3 from our dataset dimensionality reduction computational workflow. In pink, genes named as "Immunoglobulin"-encoding genes. PCA, Principal Component Analysis. PC, Principal components. ViTALs, highly variable CL-associated transcripts.

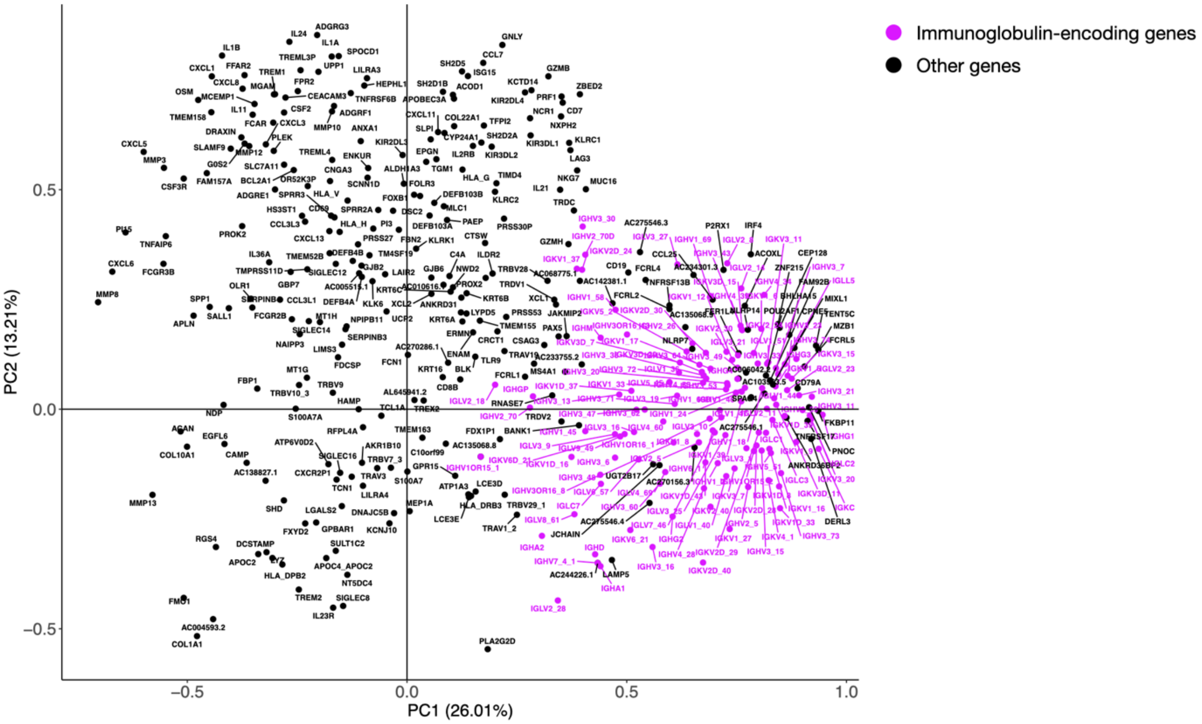

*Supplemental Figure 6. Staphylococcus isolates from CL lesions and associated clinical metadata and RNA-seq/16S-seq results*. A) 13 *S. aureus* isolates were short-read whole genome sequenced and combined with 22 publicly available *S. aureus* genomes to build out in-house *S. aureus* pangenome. A Hierarchical clustering was performed between the *S. aureus* genomes, together with a publicly available *S. epidermidis* genome to serve as comparison. B) Clinical metadata, RNA-seq and 16S-seq-associated data from the lesions where the CL S. aureus isolates originated from. C) Distribution of lesion biopsies according to the S. aureus transcript abundances (in log10 counts per million, CPM).

**A**

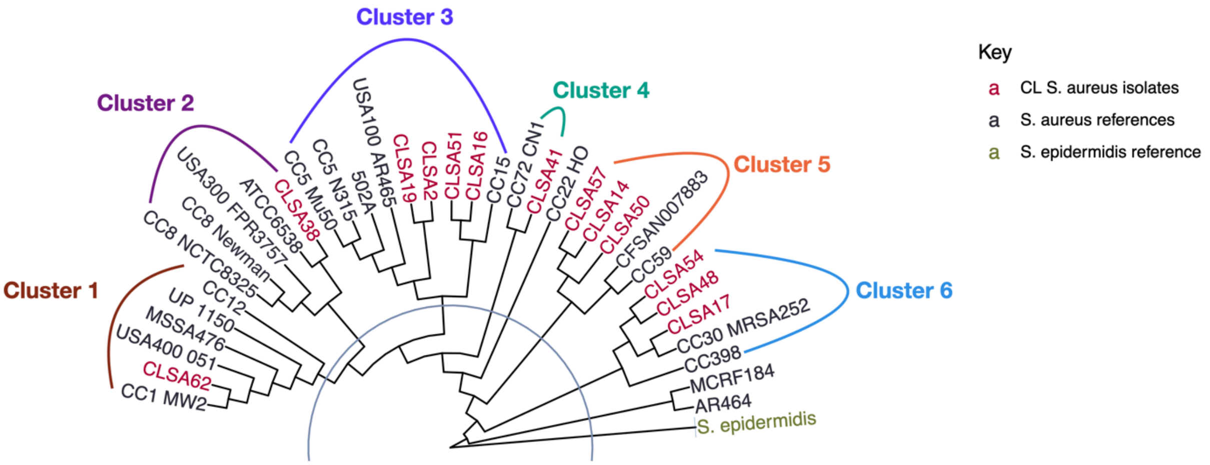


**B**

| | Clinical metadata | | | | | | | 16S-seq (swab) Microbiome profile | Lesion Biopsy | | |
|---|---|---|---|---|---|---|---|---|---|---|---|
| | | | | | | | | | RNA-seq | qPCR | |
| Cluster | CL *S. aureus* | Age | Sex | Lesion size (mm2) | Lymphadenopathy | Healing time (days) | Clinical Outcome* | Microbiome cluster | *S. aureus* (CPM) | Bacterial load | Parasite load |
| 1 | CLSA62 | 26-30 | male | 0-25th | yes | 130 | > 1 $Sb^v$ | NA | NA | NA | NA |
| 2 | CLSA38 | 41-45 | male | 50-75th | yes | 50 | 1 $Sb^v$ | NA | >75th | >75th | >75th |
| 3 | CLSA2 | 16-20 | male | 50-75th | yes | 180 | > 1 $Sb^v$ | M6 | >75th | >75th | >75th |
| 3 | CLSA16 | 16-20 | male | 50-75th | yes | 120 | > 1 $Sb^v$ | M7 | >75th | 25-50th | >75th |
| 3 | CLSA19 | 21-25 | male | 50-75th | yes | 72 | 1 $Sb^v$ | M7 | >75th | 25-50th | 50-75th |
| 3 | CLSA51 | 21-25 | male | 25-50th | yes | 64 | 1 $Sb^v$ | NA | 50-75th | 25-50th | 50-75th |
| 4 | CLSA41 | 31-35 | male | >75th | no | 120 | > 1 $Sb^v$ | NA | >75th | >75th | 25-50th |
| 5 | CLSA14 | 46-50 | male | >75th | yes | 221 | > 1 $Sb^v$ | M6 | 25-50th | >75th | >75th |
| 5 | CLSA50 | 16-20 | male | 0-25th | yes | 63 | 1 $Sb^v$ | NA | >75th | 0-25th | 50-75th |
| 5 | CLSA57 | 31-35 | male | 25-50th | yes | 180 | > 1 $Sb^v$ | M6 | NA | NA | NA |
| 6 | CLSA17 | 21-25 | female | 25-50th | no | 45 | 1 $Sb^v$ | M6 | 25-50th | 50-75th | 0-25th |
| 6 | CLSA48 | 36-40 | female | 25-50th | no | 100 | > 1 $Sb^v$ | NA | >75th | 0-25th | >75th |
| 6 | CLSA54 | 16-20 | male | 0-25th | yes | 135 | > 1 $Sb^v$ | M6 | NA | NA | NA |

Picked for exp. in mice → CLSA50

For continuous variables:
- >75th quartile
- 50-75th quartile
- 25-50th quartile
- 0-25th quartile

Relative to values in the entire cohort of 62 patients

*1 round of antimony $Sb^v$ = Cure, >1 round of antimony $Sb^v$ = Failure

C

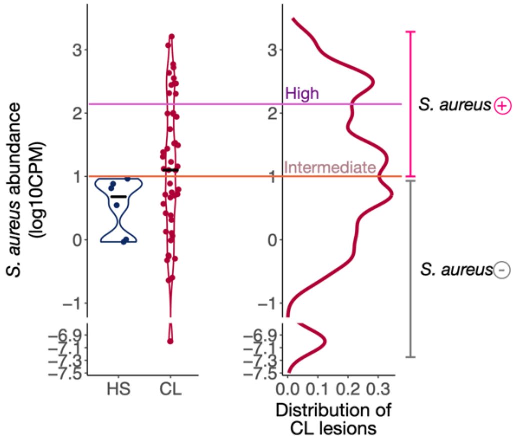

S. aureus abundance (log10CPM)
3
2
1
0
−1
−6.9
−7.1
−7.3
−7.5
HS
CL
High
Intermediate
0.0 0.1 0.2 0.3
Distribution of CL lesions
S. aureus +
S. aureus -

*Supplemental Figure 7. Number of CD90.2-, αβT cells and CD4+ T cells in S. aureus and L. braziliensis treatment protocol in C57BL/6 mice, with blockade of IL-1α, IL-1β and IL-1R with monoclonal antibodies.* Number of CD90.2-, αβT cells and CD4+ T cells recovered from the co-infected ears in week six. Non-parametric Mann-Whitney test was used for statistical significance, *P<0.05, **P<0.01, ***P<0.001. Violin plots and median are represented in the plots when appropriate.

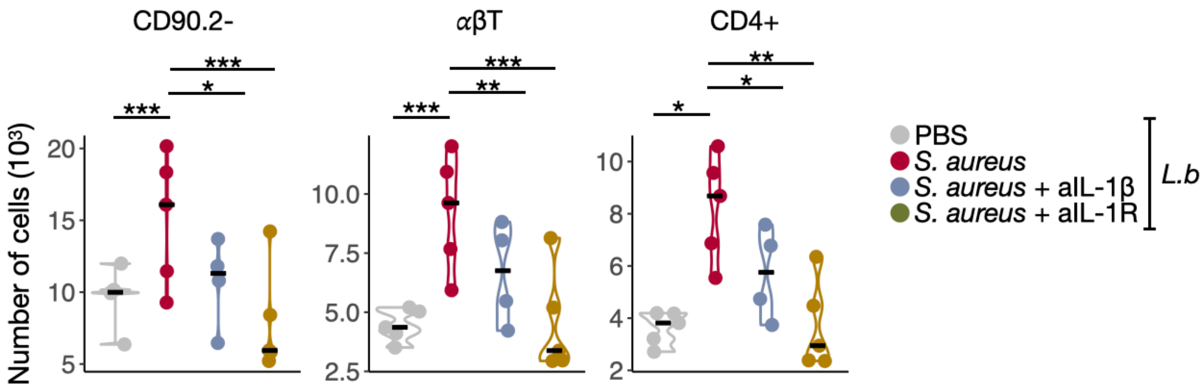
